# Harmonising APASL and A-TANGO criteria for acute-on-chronic liver failure: identification of complementary high-risk pre-ACLF populations

**DOI:** 10.64898/2026.05.22.26353839

**Authors:** Nipun Verma, Pratibha Garg, Gowri Priya Nair, Arathi Venu, Nikhil Sai Jarpula, Parminder Kaur, Arka De, Madhumita Premkumar, Sunil Taneja, Tarana Gupta, Arun Valsan, Ajay Duseja, Rajiv Jalan

## Abstract

**Background & Aims:** ACLF is defined differently by APASL (acute hepatic dysfunction) and by organ failure-based frameworks including EASL-CLIF and the recently developed A-TANGO score. Whether these definitions identify competing populations or sequential stages of the same syndrome remains unresolved, with direct implications for the timing of intervention. We tested whether APASL-defined ACLF can be integrated into the A-TANGO framework to identify a clinically actionable patient population.

**Methods:** 4,024 patients hospitalised with acute decompensation of cirrhosis in a multicentre cohort were classified simultaneously by APASL and A-TANGO criteria. Mortality, progression to A-TANGO ACLF among A-TANGO-negative patients, and reversal of ACLF were assessed using Fine-Gray competing-risk models with death as a competing event. EASL-CLIF analyses were performed as sensitivity analyses.

**Results:** A-TANGO-negative/APASL-positive patients comprised 8.7% of the cohort and had higher 90-day mortality than A-TANGO-negative/APASL-negative patients (22.3% vs 14.4%, p=0.001), despite similar 28-day mortality. Once A-TANGO ACLF was established, 28-day mortality was high irrespective of APASL status (45.4% in APASL-positive and 56.0% in APASL-negative patients). Among A-TANGO-negative patients, 53.5% of APASL-positive vs 27.9% of APASL-negative patients progressed to A-TANGO ACLF within 28 days, with APASL positivity independently predicting progression (adjusted sHR: 2.30, 95%CI: 1.90-2.77). Within A-TANGO-negative/APASL-negative patients an A-TANGO OF score ≥8 independently enriched for progression (52% vs 19%). A-TANGO reversal occurred in 17.1% and was independently reduced by APASL positivity (adjusted sHR: 0.756, 95%CI: 0.586-0.975), while APASL reversal was rare (4.0%). EASL-CLIF sensitivity analyses were directionally consistent.

**Conclusions:** APASL-defined ACLF does not compete with A-TANGO; it occupies an upstream position on the same disease trajectory. A-TANGO-negative/APASL-positive patients and A-TANGO-negative/APASL-negative patients with A-TANGO OF ≥8 represent complementary pre-ACLF populations suitable for prevention trials and enrichment strategies.

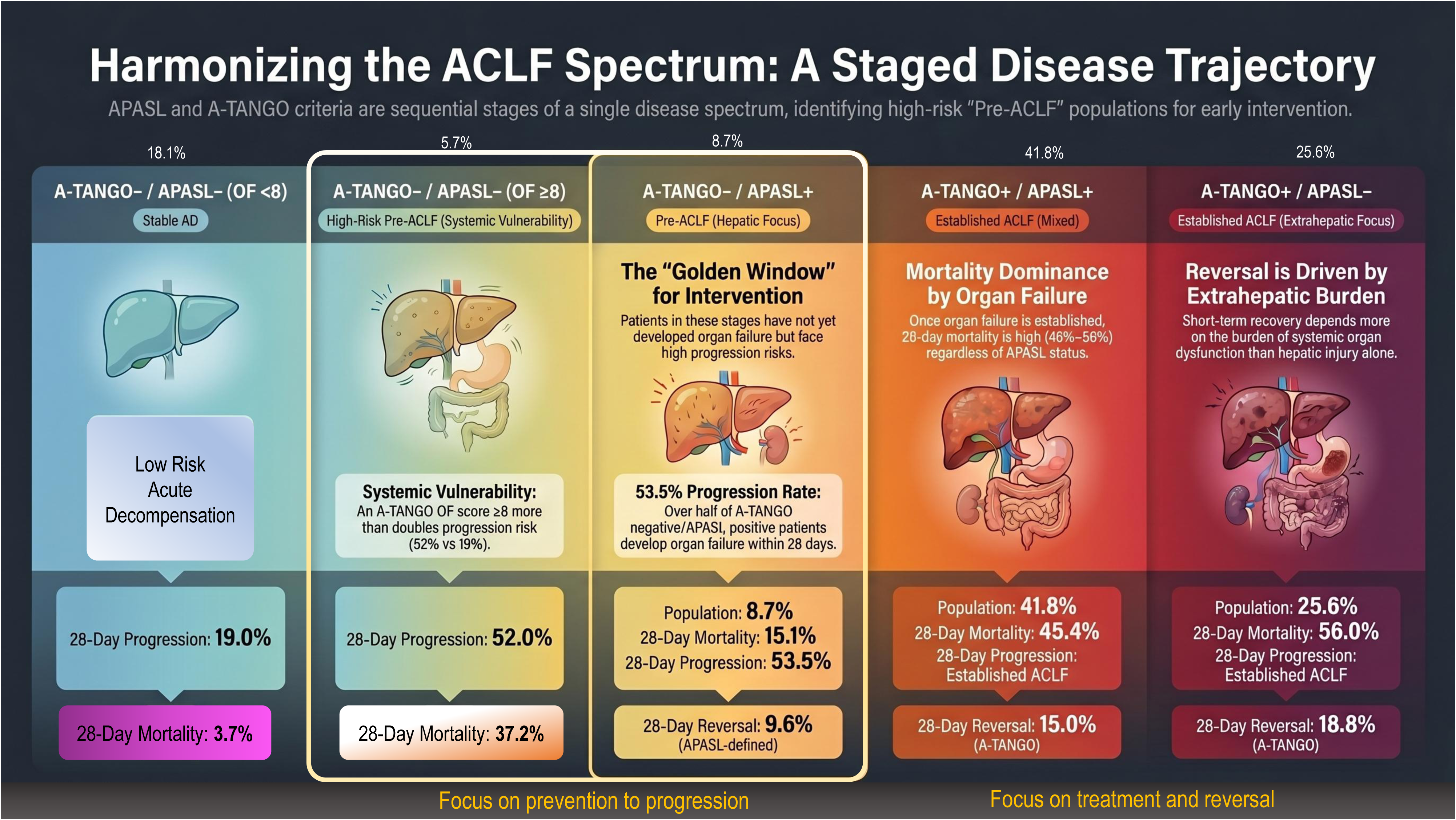

## INTRODUCTION

Acute-on-chronic liver failure (ACLF) is a dynamic syndrome of acute deterioration in patients with chronic liver disease, characterised by intense systemic inflammation, rapidly evolving organ dysfunction and high short-term mortality(1). Over the past decade three competing diagnostic frameworks have shaped clinical practice and the design of clinical trials(2–4). The EASL-CLIF criteria define ACLF on the basis of extrahepatic organ failure assessed by the CLIF-C Organ Failure score(2). The APASL criteria define ACLF on the basis of acute hepatic dysfunction, specifically jaundice and coagulopathy followed within four weeks by ascites or hepatic encephalopathy (4, 5). The recently developed A-TANGO organ failure score recalibrates organ failure thresholds against contemporary mortality risk in large global cohorts and refines the CLIF-C OF framework for current practice(6). Continuing with the efforts, a global panel of experts described a consensus definition of organ failures and ACLF, where “liver failure” characterised by bilirubin>=7.5 and INR>= 1.5 with an extra-hepatic organ failure was proposed as ACLF(7). Subsequently, this framework was compared with A-TANGO criteria in two large multi-national cohorts and was identified to have a lower sensitivity with a considerable proportion of patients in no-ACLF group that were deemed high risk ACLF by the A-TANGO criteria(8). Although each definition has independent prognostic validity, they identify partially overlapping populations and have been treated, sometimes unhelpfully, as competing rather than complementary descriptions of the same disease. A recent paper confirmed this speculation demonstrating that the A-TANGO and Chinese Group on the Study of Severe Hepatitis B-ACLF frameworks are complimentary than competitive(9).

The unresolved heterogeneity between the ACLF definitions has practical consequences. Discordant criteria produce inconsistent estimates of disease burden, lead to variable patient selection in clinical trials, and create uncertainty about the optimal timing of intervention(9–11). It is becoming clear that these frameworks may not simply disagree on thresholds; they may capture sequential stages of the same biological process(10). APASL identifies patients with intense acute hepatic injury that has not yet declared itself as multi-organ failure(4). A-TANGO and EASL-CLIF identify patients in whom organ failure has already developed(2, 6). The Kyoto APASL Consensus acknowledged this explicitly and proposed that future definitions should accommodate both hepatic dysfunction-dominant and organ failure-dominant phases of the syndrome(5). Whether the two frameworks describe distinct entities or adjacent stages within a single ACLF continuum remains the central unresolved question in the field.

Resolving this question matters because it determines where on the disease trajectory we should intervene. Organ failure-based criteria, including A-TANGO and EASL-CLIF, are highly accurate for prognostication once ACLF is established, but are by definition limited to a late stage of the syndrome(2, 6). A framework that identifies the patient at the cusp of organ failure, in whom the trajectory is still modifiable, would address one of the major unmet needs in decompensated cirrhosis.

In this study, we tested whether APASL-defined hepatic dysfunction can be integrated into the A-TANGO organ failure framework in a large multicentre cohort of patients hospitalised with acute decompensation of cirrhosis. We addressed three specific questions. First, what do the two scores capture in terms of clinical profile and short-term mortality when applied simultaneously. Second, does APASL positivity identify patients without baseline organ failure who progress to A-TANGO ACLF. Third, does APASL positivity at baseline modify the probability of reversal of ACLF once organ failure is established. EASL-CLIF analyses were used as sensitivity analyses to confirm consistency across organ failure frameworks.

## METHODS

### Study design, setting, and population

This ambispective multicentre cohort study included adults hospitalized with acute decompensation of cirrhosis at three tertiary care centres in India: Postgraduate Institute of Medical Education and Research, Chandigarh; Amrita Institute of Medical Sciences, Kochi; and Post Graduate Institute of Medical Sciences, Rohtak. Patients were enrolled between 2015 and 2023 and study was approved by the ethics committee (PGI/IEC-08/2021-2062).

Eligible patients were aged ≥18 years and had cirrhosis with acute decompensation, defined according to EASL criteria(2). Patients were excluded if they had hepatocellular carcinoma or another active malignancy, previous hepatic or extrahepatic organ transplantation, HIV infection, pregnancy or lactation, missing variables required to classify ACLF by the study definitions, or refusal of consent. Patients were followed for up to 90 days from admission and managed according to standard of care, including intensive care support when clinically indicated. No patient underwent liver transplantation during follow-up. The study was approved by the Institutional Ethics Committees of the participating centres. Written informed consent was obtained from prospectively enrolled patients, and a waiver of consent was granted for retrospectively included cases.

### ACLF definitions and baseline classification

ACLF was classified at admission using APASL, A-TANGO, and EASL-CLIF criteria. The primary analysis used A-TANGO as the organ-failure ACLF framework, with APASL used to define patients with acute hepatic dysfunction. EASL-CLIF was used for sensitivity analyses.

APASL ACLF was defined as acute hepatic dysfunction in a patient with previously diagnosed or undiagnosed chronic liver disease, characterized by jaundice with serum bilirubin ≥5 mg/dL and coagulopathy with INR ≥1.5, complicated within 4 weeks by ascites and/or hepatic encephalopathy. As per recent updates, the APASL criteria were applied irrespective of whether the precipitating event was hepatic or extrahepatic in origin(5).

A-TANGO organ failure(6) was defined by the presence of a subscore of 3 in the respective organ system. The thresholds were: serum bilirubin ≥20 mg/dL for liver failure; serum creatinine ≥2 mg/dL, acute kidney injury stage 1b, or requirement for renal replacement therapy for kidney failure; West Haven grade III–IV hepatic encephalopathy or need for mechanical ventilation for cerebral failure; INR ≥2.2 for coagulation failure; requirement for vasopressor support for circulatory failure; and SpO_₂_/FiO_₂_ ≤325 or requirement for mechanical ventilation for respiratory failure. A-TANGO ACLF(6) was diagnosed in the presence of at least one liver, kidney, coagulation, or respiratory failure. Isolated cerebral or circulatory failure was considered A-TANGO ACLF only when accompanied by an additional organ dysfunction. Patients fulfilling these criteria were classified as A-TANGO+, and those not fulfilling them were classified as A-TANGO−.

EASL-CLIF ACLF(2) was defined using CLIF-C Organ Failure score thresholds. Liver failure was defined as serum bilirubin ≥12 mg/dL; kidney failure as serum creatinine ≥2 mg/dL or requirement for renal replacement therapy; cerebral failure as West Haven grade III–IV hepatic encephalopathy; coagulation failure as INR ≥2.5; circulatory failure as requirement for vasopressor support; and respiratory failure as SpO_₂_/FiO_₂_ ≤214 or requirement for mechanical ventilation. EASL-CLIF ACLF was diagnosed according to the number and type of organ failures and associated renal or cerebral dysfunction, as previously described.

### Harmonized ACLF phenotypes

To examine the relationship between hepatic dysfunction-based and organ failure-based ACLF frameworks, patients were stratified into four mutually exclusive baseline phenotypes according to APASL and A-TANGO status, a) A-TANGO-negative/APASL-positive representing acute decompensation without ACLF by either framework; b) A-TANGO-negative/APASL-positive, representing APASL-defined acute hepatic dysfunction without A-TANGO ACLF; c) A-TANGO-positive/APASL-negative, representing A-TANGO organ failure-based ACLF without APASL-defined acute hepatic dysfunction; and d) A-TANGO-positive/APASL-positive, representing overlap between APASL-defined acute hepatic dysfunction and A-TANGO ACLF. This classification allowed us to determine whether APASL identifies a clinically relevant phenotype within patients who do not yet meet A-TANGO ACLF criteria.

### Baseline description and clinical outcomes

Baseline characteristics, ACLF phenotype distribution, and short-term outcomes were first described across the four A-TANGO/APASL groups. Outcomes included 28-day and 90-day mortality. Survival was estimated using Kaplan–Meier methods and compared using the log-rank test. Cox proportional hazards models were used to estimate hazard ratios for 28-day mortality across baseline phenotypes, using A-TANGO-negative/APASL-negative as the reference group.

### Progression to ACLF

Progression analyses were restricted to patients who were A-TANGO-negative at admission. Progression was defined as development of A-TANGO ACLF within 28 days after admission. Patients were compared according to APASL status at baseline, specifically A-TANGO-negative/APASL-negative versus A-TANGO-negative/APASL-positive. Because death before ACLF onset precludes subsequent observation of progression, the cumulative incidence of progression to A-TANGO ACLF over 28 days was estimated using competing-risk methods, with death treated as a competing event. Liver transplantation was not included as a competing event because no patient underwent transplantation during follow-up. Groups were compared using Gray’s test.

### Predictors of progression

Predictors of 28-day progression to A-TANGO ACLF were evaluated among patients without A-TANGO ACLF at admission. Candidate variables included APASL status, baseline A-TANGO organ failure score, infection, markers of hepatic and renal dysfunction, systemic inflammation, and other clinically relevant variables. Variables were selected for multivariable modelling based on clinical relevance and univariable associations, while avoiding collinearity (variance inflation factor <5) between component variables and composite severity scores. Subdistribution hazard ratios were estimated using Fine–Gray competing-risk regression models, with death treated as a competing event. Multivariable models were adjusted for age, sex, and etiology of cirrhosis, with additional covariates included according to the model structure.

Additional analyses were performed within the A-TANGO-negative/APASL-negative subgroup to identify patients at increased risk of subsequent A-TANGO ACLF despite not fulfilling either ACLF definition at admission. A-TANGO OF score thresholds were evaluated for discrimination using receiver operating characteristic analysis. Optimal cut-off was identified using the Youden index.

### Reversal of ACLF

Reversal of ACLF was assessed at the last available evaluation before death, discharge, or day 28, whichever occurred first. A-TANGO reversal was evaluated among patients with A-TANGO ACLF at admission, including both A-TANGO-positive/APASL-negative and A-TANGO-positive/APASL-positive phenotypes. A-TANGO reversal was defined as no longer fulfilling A-TANGO ACLF criteria. Because death before reversal precludes subsequent observation of recovery, the 28-day cumulative incidence of A-TANGO reversal was estimated using competing-risk methods, with death treated as a competing event. Groups were compared using Gray’s test. APASL reversal was evaluated among APASL+ patients and was defined as resolution of acute hepatic dysfunction, indicated by serum bilirubin <5 mg/dL, INR <1.5, and resolution or control of ascites and hepatic encephalopathy.

### Predictors of reversal

Predictors of A-TANGO reversal were evaluated among patients with A-TANGO ACLF at admission. Candidate variables included APASL status, A-TANGO OF score, individual organ failures, laboratory parameters, infection, and baseline disease severity. Regression models were adjusted for age, sex, and etiology of cirrhosis where appropriate. Competing-risk estimates for reversal were generated because death before reversal prevents subsequent observation of recovery.

### Sensitivity analysis using EASL-CLIF ACLF

All key analyses were repeated using EASL-CLIF ACLF in place of A-TANGO ACLF to assess consistency across organ-failure frameworks. Patients were reclassified into EASL-negative/APASL-negative, EASL-negative/APASL-positive, EASL-positive/APASL-negative, and EASL-positive/APASL-positive phenotypes. Baseline phenotype distribution, 28-day and 90-day mortality, progression to EASL-CLIF ACLF among EASL-negative patients, and reversal among EASL-positive patients were evaluated using the same analytical approach.

### Statistical analysis

Continuous variables are presented as median with interquartile range and were compared using the Mann–Whitney U test or Kruskal–Wallis test, as appropriate. Categorical variables are presented as counts and percentages and were compared using the chi-square test or Fisher’s exact test. Survival was estimated using Kaplan-Meier analysis and compared using the log-rank test. Cox proportional hazards models were used to estimate hazard ratios for 28-day mortality. For 28-day ACLF progression and reversal, cumulative incidence functions were estimated using competing-risk methods, with death treated as a competing event. Fine–Gray regression was used to estimate subdistribution hazard ratios for progression. All analyses were performed using SPSS and RStudio. A two-sided p value <0.05 was considered statistically significant.

## RESULTS

### Study population and baseline characteristics

A total of 4,024 patients hospitalized with acute decompensation of cirrhosis were included. The median age was 45 years, and 89.1% were male. Alcohol-associated liver disease was the predominant etiology of cirrhosis, followed by viral hepatitis and MASLD. Alcohol-associated hepatitis, either alone or in combination with another precipitant, was the most frequent acute insult, and hepatic precipitants were more common than extrahepatic precipitants. Ascites and hepatic encephalopathy were frequent at presentation, reflecting an advanced decompensated cohort. No patient underwent liver transplantation during follow-up **Table S1.**

### Distribution of harmonized A-TANGO/APASL phenotypes

Using the harmonized A-TANGO/APASL framework, patients were classified into four mutually exclusive admission phenotypes **(Figure 1 and Table 1)**: A-TANGO-negative/APASL-negative in 959 patients (23.8%), A-TANGO-negative/APASL-positive in 350 patients (8.7%), A-TANGO-positive/APASL-positive in 1,683 patients (41.8%), and A-TANGO-positive/APASL-negative in 1,032 patients (25.6%). Thus, APASL-alone disease represented a small subgroup, whereas most APASL-positive patients were already captured within the A-TANGO ACLF framework.

**Figure 1:**
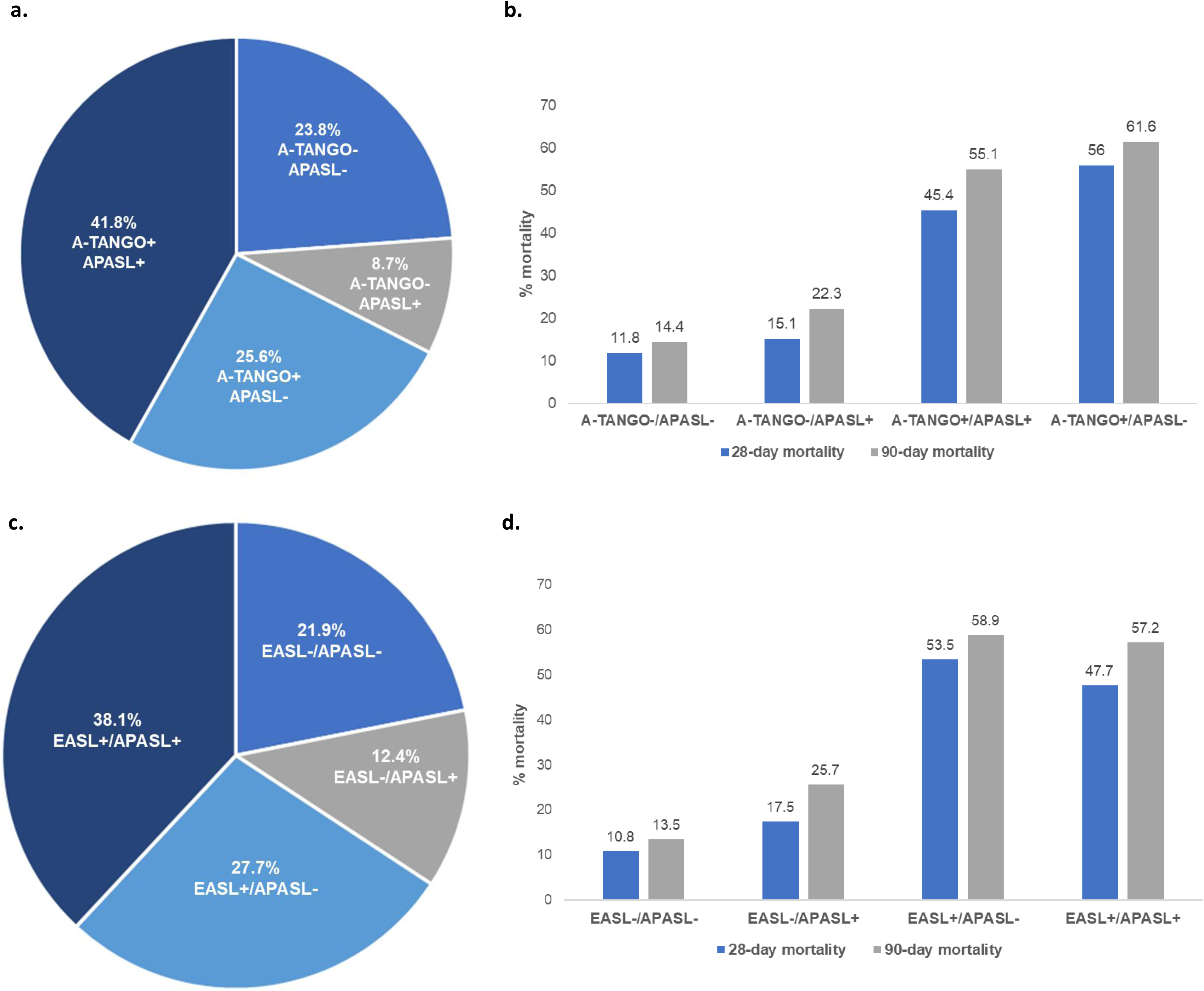
Distribution of harmonized ACLF phenotypes and associated mortality outcomes. (a) Distribution of patients across A-TANGO/APASL phenotypes within the overall cohort. (b) Twenty-eight-day and 90-day transplant-free mortality across A-TANGO/APASL phenotypes. (c) Distribution of patients across EASL/APASL phenotypes within the overall cohort. (d) Twenty-eight-day and 90-day transplant-free mortality across EASL/APASL phenotypes.

**Table 1:** Characteristics of acutely decompensated cohort stratified by APASL criteria.

|  | A-TANGO- |  |  | A-TANGO+ |  |  |  |
| --- | --- | --- | --- | --- | --- | --- | --- |
| Characteristic | APASL-<br>(n= 959,<br>23.8%) | APASL+<br>(n= 350,<br>8.7%) | P-<br>value | APASL+<br>(n= 1683,<br>41.8%) | APASL-<br>(n= 1032,<br>25.6%) | P-<br>value | Overall<br>p-<br>value |
| Age, median<br>(IQR) | 47.0<br>(41.0 -<br>55.0) | 45.0<br>(39.0 -<br>53.0) | 0.019 | 45.0<br>(36.0 -<br>53.0) | 46.0 (38.0<br>- 55.0) | <0.001 | <0.001 |
| Female sex, n/N<br>(%) | 101<br>(10.5) | 37 (10.6) | 1.000 | 194<br>(11.5) | 108 (10.5) | 0.429 | 0.790 |
| **Etiology of cirrhosis (n, %)** |  |  |  |  |  |  |  |
| Alcohol | 711<br>(74.1) | 255<br>(72.9) | 0.004 | 1178<br>(70.0) | 729 (70.6) | <0.001 | <0.001 |
| MASLD | 73 (7.6) | 12 (3.4) |  | 112 (6.7) | 111 (10.8) |  |  |
| Hepatitis B/C | 71 (7.4) | 44 (12.6) |  | 200<br>(11.9) | 48 (4.7) |  |  |
| Alcohol+ CMRF | 22 (2.3) | 3 (0.9) |  | 17 (1.0) | 27 (2.6) |  |  |
| AIH | 33 (3.4) | 13 (3.7) |  | 78 (4.6) | 67 (6.5) |  |  |
| Cryptogenic | 25 (2.6) | 13 (3.7) |  | 72 (4.3) | 11 (1.1) |  |  |
| Alcohol+ Hepatitis<br>B/C | 20 (2.1) | 10 (2.9) |  | 22 (1.3) | 34 (3.3) |  |  |
| Others | 4 (0.4) | 0 (0.0) |  | 4 (0.2) | 5 (0.5) |  |  |
| **Precipitating events (n, %)** |  |  |  |  |  |  |  |
| AH | 110<br>(11.5) | 157<br>(44.9) | <0.001 | 688<br>(40.9) | 164 (15.9) | <0.001 | <0.001 |
| Infections | 143<br>(14.9) | 35 (10.0) |  | 199<br>(11.8) | 299 (29.0) |  |  |
| AH+ others | 242<br>(25.2) | 67 (19.1) |  | 335<br>(19.9) | 387 (37.5) |  |  |
| DILI | 36 (3.8) | 21 (6.0) |  | 131 (7.8) | 38 (3.7) |  |  |
| Viral hepatitis | 9 (0.9) | 42 (12.0) |  | 171<br>(10.2) | 28 (2.7) |  |  |
| AIH | 18 (1.9) | 2 (0.6) |  | 29 (1.7) | 26 (2.5) |  |  |
| UGIB/LGIB | 377<br>(39.3) | 13 (3.7) |  | 22 (1.3) | 63 (6.1) |  |  |
| Others | 24 (2.5) | 13 (3.7) |  | 108 (6.4) | 27 (2.6) |  |  |
| **Type of acute insult** |  |  |  |  |  |  |  |
| Hepatic | 415<br>(43.3) | 289<br>(82.6) | <0.001 | 1354<br>(80.5) | 643 (62.3) | <0.001 | <0.001 |
| Extra-hepatic | 544<br>(56.7) | 61 (17.4) |  | 329<br>(19.5) | 389 (37.7) |  |  |
| More than acute<br>insult | 459<br>(47.9) | 90 (25.7) | <0.001 | 449<br>(26.7) | 437 (42.3) | <0.001 | <0.001 |
| **Prior**<br>**decompensation** | 117<br>(29.5) | - | - | - | 465 (96.1) | - | <0.001 |
| **Decompensations** |  |  |  |  |  |  |  |
| **Prevalence of** | 741 | 328 | <0.001 | 1657 | 969 (93.9) | <0.001 | <0.001 |
| **Ascites** | (77.3) | (93.7) |  | (98.5) |  |  |  |
| **Ascites grade** |  |  |  |  |  |  |  |
| No Ascites | 34 (5.6) | 22 (6.5) | <0.001 | 26 (1.6) | 54 (5.4) | <0.001 | <0.001 |
| Mild to moderate | 343 (56.7) | 249 (73.2) |  | 1245 (74.2) | 519 (51.6) |  |  |
| Tense ascites | 228 (37.7) | 69 (20.3) |  | 406 (24.2) | 433 (43.0) |  |  |
| **Prevalence of HE** | 481 (50.2) | 186 (53.1) | 0.371 | 1050 (62.4) | 807 (78.2) | <0.001 | <0.001 |
| **HE grades** |  |  |  |  |  |  |  |
| No HE | 169 (27.9) | 174 (51.2) | <0.001 | 630 (37.6) | 208 (20.7) | <0.001 | <0.001 |
| Grade I-II | 429 (70.9) | 166 (48.8) |  | 857 (51.1) | 549 (54.6) |  |  |
| Grade III-IV | 7 (1.2) | 0 (0.0) |  | 190 (11.3) | 249 (24.8) |  |  |
| **Infection at enrolment** | 133 (22.0) | 79 (23.2) | 0.718 | 586 (34.9) | 517 (51.4) | <0.001 | <0.001 |
| **Lab investigations** |  |  |  |  |  |  |  |
| Haemoglobin (gm/dL) | 8.8 (7.2 - 10.7) | 10.2 (8.4 - 11.8) | <0.001 | 10.0 (8.4 - 11.5) | 8.7 (7.2 - 10.4) | <0.001 | <0.001 |
| WBCC (x10<sup>9</sup>/L) | 7.8 (5.5 - 10.9) | 8.3 (6.2 - 12.0) | 0.049 | 11.7 (7.4 - 17.1) | 11.2 (6.8 - 16.7) | 0.096 | <0.001 |
| Platelet count (x10<sup>9</sup>/L) | 91.0 (67.0 - 132.0) | 105.0 (67.0 - 159.0) | 0.004 | 108.0 (68.0 - 171.5) | 88.5 (57.0 - 138.2) | <0.001 | <0.001 |
| INR | 1.3 (0.9 - 1.6) | 1.8 (1.6 - 2.0) | <0.001 | 2.3 (1.9 - 2.9) | 2.0 (1.4 - 2.6) | <0.001 | <0.001 |
| AST (U/L) | 81.0 (53.0 - 134.0) | 116.0 (78.0 - 188.0) | <0.001 | 140.0 (90.3 - 214.0) | 94.0 (59.7 - 154.0) | <0.001 | <0.001 |
| ALT (U/L) | 41.0 (25.0 - 66.4) | 53.4 (31.9 - 90.5) | <0.001 | 58.0 (35.0 - 113.0) | 42.0 (27.5 - 73.9) | <0.001 | <0.001 |
| Bilirubin (mg/dL) | 3.2 (1.5 - 6.8) | 9.5 (6.9 - 14.7) | <0.001 | 20.9 (12.2 - 27.2) | 13.0 (4.3 - 23.7) | <0.001 | <0.001 |
| Creatinine (mg/dL) | 0.9 (0.7 - 1.1) | 0.9 (0.7 - 1.2) | 0.979 | 1.3 (0.8 - 2.4) | 1.5 (0.9 - 2.6) | <0.001 | <0.001 |
| Sodium (mmol/L) | 134.0 (130.0 - 137.0) | 132.7 (129.0 - 136.0) | 0.002 | 131.0 (126.7 - 135.0) | 132.0 (128.0 - 137.0) | <0.001 | <0.001 |
| MAP (mmHg) | 81.7 (73.3 - 89.7) | 85.0 (78.0 - 92.5) | <0.001 | 84.0 (77.3 - 92.0) | 85.0 (77.0 - 93.3) | <0.001 | <0.001 |
| **Disease severity scores** |  |  |  |  |  |  |  |
| MELD score | 15.0 (12.0 - 17.6) | 22.5 (20.9 - 24.4) | <0.001 | 30.5 (26.5 - 36.5) | 27.2 (21.8 - 32.8) | <0.001 | <0.001 |
| MELD-Na score | 19.4 | 25.7 | <0.001 | 32.7 | 29.7 (25.2 | <0.001 | <0.001 |
|  | (15.7 - 22.9) | (22.6 - 27.9) |  | (29.2 - 37.4) | - 34.5) |  |  |
| ATANGO- OF score | 7.0 (6.0 - 7.0) | 8.0 (8.0 - 8.0) | <0.001 | 11.0 (10.0 - 13.0) | 11.0 (9.0 - 13.0) | <0.001 | <0.001 |
| AARC score | 8.0 (7.0 - 8.0) | 8.0 (7.0 - 9.0) | 0.051 | 10.0 (8.0 - 11.0) | 10.0 (9.0 - 12.0) | 0.297 | <0.001 |
| **Organ failures as per A-TANGO criteria** |  |  |  |  |  |  |  |
| Cerebral failure | 7 (0.7) | 0 (0.0) | 0.240 | 190 (11.3) | 249 (24.1) | <0.001 | <0.001 |
| Coagulation failure | 0 (0.0) | 0 (0.0) | - | 1012 (60.1) | 457 (44.3) | <0.001 | <0.001 |
| Liver failure | 0 (0.0) | 0 (0.0) | - | 912 (54.2) | 371 (35.9) | <0.001 | <0.001 |
| Kidney failure | 0 (0.0) | 0 (0.0) | - | 582 (34.6) | 406 (39.3) | 0.014 | <0.001 |
| Circulatory failure | 11 (1.1) | 1 (0.3) | 0.263 | 523 (31.1) | 353 (34.2) | 0.099 | <0.001 |
| Respiratory failure | 0 (0.0) | 0 (0.0) | - | 504 (29.9) | 237 (23.0) | <0.001 | <0.001 |
| **28-day mortality** | 113 (11.8) | 53 (15.1) | 0.128 | 764 (45.4) | 578 (56.0) | <0.001 | <0.001 |
| **90-day mortality** | 138 (14.4) | 78 (22.3) | 0.001 | 928 (55.1) | 636 (61.6) | <0.001 | <0.001 |
| **Progression to A-TANGO ACLF over 28-days\*** | 169/605 (27.9) | 182/340 (53.5) | <0.001 | - | - | - | - |
| **Reversal of ACLF as per ATANGO\*** | - | - | - | 90/602 (15.0) | 148/786 (18.8) | 0.067 | - |
| **Reversal of ACLF as per APASL\*** | - | 15/156 (9.6) | - | 16/612 (2.6) | - | - | <0.001 |

INR: international normalized ration, AST: Aspartate Aminotransferase, ALT: Aspartate Aminotransferase, MAP: Mean arterial pressure, MELD: Model of end-stage liver disease, MELD-Na: MELD-sodium
Categorical variables are presented as n (%), and continuous variables as median (interquartile range, IQR). Categorical variables were compared using the Chi-square test, and continuous variables using the Mann–Whitney U test. A two-sided p-value <0.05 was considered statistically significant.
\*Percentages are calculated among participants with available data.

### APASL-defined hepatic dysfunction identifies a distinct phenotype within A-TANGO strata

Within the A-TANGO-negative population, APASL-positive patients had a distinct hepatic dysfunction phenotype compared with A-TANGO-negative/APASL-negative patients (**Table 1).** They had higher bilirubin, INR, aminotransferases, MELD and MELD-Na scores, and were more likely to have hepatic precipitants, particularly alcohol-associated hepatitis and viral hepatitis-related insults. Despite not meeting A-TANGO ACLF criteria at admission, this group had 28-day mortality not different than A-TANGO-negative/APASL-negative patients, although 90-day mortality was statistically higher in APASL-positive cases.

Among patients with A-TANGO ACLF at admission, APASL status continued to distinguish clinical phenotype **(Table 1).** A-TANGO-positive/APASL-positive patients had greater hepatic dysfunction, with higher bilirubin and INR and more frequent hepatic precipitants. In contrast, A-TANGO-positive/APASL-negative patients had greater extrahepatic/systemic involvement, including more infection, advanced hepatic encephalopathy, cerebral failure, kidney failure, circulatory failure, and respiratory failure. However, once A-TANGO ACLF was present, both APASL-positive and APASL-negative phenotypes had very high short-term mortality, indicating that established organ failure dominated prognosis.

Kaplan-Meier analysis **(Figure 2)** showed clear separation in survival across the four phenotypes. In Cox regression, A-TANGO-negative/APASL-positive patients had a 28-day mortality risk not different than A-TANGO-negative/APASL-negative patients. In contrast, both A-TANGO-positive phenotypes had markedly increased mortality risk, with the highest risk in A-TANGO-positive/APASL-negative patients, followed by A-TANGO-positive/APASL-positive patients **(Table S2).** Together, these findings support a clinical continuum in which A-TANGO-negative/APASL-positive represents an intermediate hepatic dysfunction state between acute decompensation without ACLF and established A-TANGO organ failure-defined ACLF.

**Figure 2:**
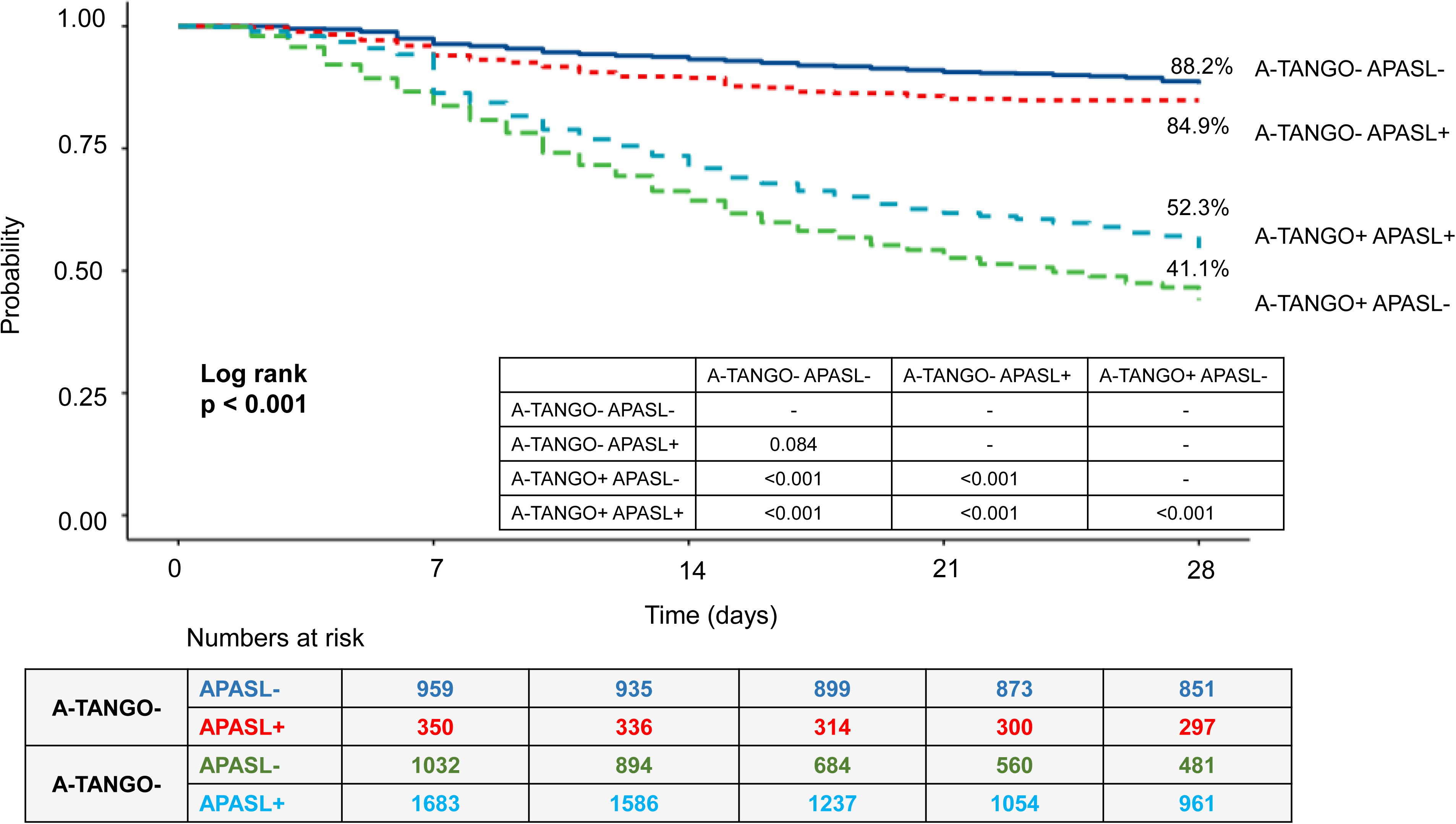
Kaplan-Meier survival analysis according to harmonized ACLF phenotypes. Kaplan-Meier curves demonstrating 28-day transplant-free survival stratified according to combined A-TANGO and APASL phenotypes. Survival differences between groups were assessed using the log-rank test. Pairwise comparisons between phenotypes are displayed within the figure. Numbers at risk are shown below the curves. A two-sided p-value <0.05 was considered statistically significant.

### Progression to A-TANGO ACLF among patients without baseline A-TANGO ACLF

Progression analyses were restricted to patients who were A-TANGO-negative at admission. Among patients with available follow-up, 351 of 945 patients (37.1%) progressed to A-TANGO ACLF within 28 days. Progression was substantially more frequent in A-TANGO-negative/APASL-positive patients than in A-TANGO-negative/APASL-negative patients: 182 of 340 patients (53.5%) versus 169 of 605 patients (27.9%), respectively **(Figure 3).** Thus, more than half of APASL-alone patients progressed to organ failure-defined ACLF within 28 days.

**Figure 3:**
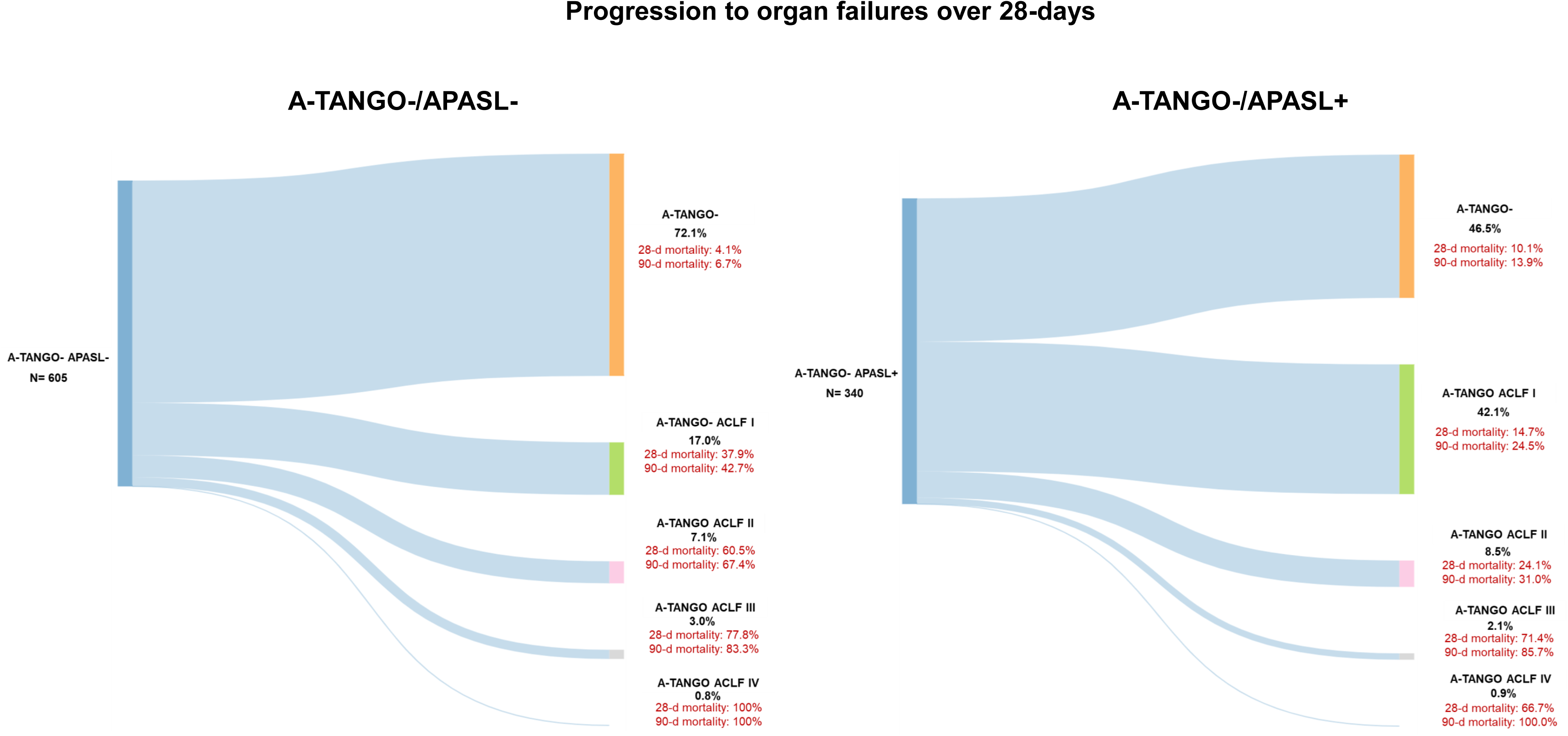
Longitudinal progression to organ failure over 28 days among patients without baseline A-TANGO ACLF. Sankey plots illustrating disease trajectories among patients classified as A-TANGO−/APASL− and A-TANGO−/APASL+ at baseline. Transitions from baseline non-ACLF states to subsequent A-TANGO ACLF grades over 28 days are shown, together with corresponding 28-day and 90-day mortality within each trajectory. Percentages represent proportions of patients progressing to each disease state.

Patients who progressed had greater baseline disease severity, including higher bilirubin, INR, MELD, MELD-Na, A-TANGO OF, and AARC scores, lower sodium, higher leukocyte count, and more frequent infection and hepatic encephalopathy. Progression was clinically meaningful: 28-day mortality was 33.9% among progressors compared with 5.7% among non-progressors, and 90-day mortality was 41.6% versus 8.6%, respectively **(Table S3).**

Competing-risk analysis, with death treated as a competing event, confirmed a higher cumulative incidence of A-TANGO ACLF onset among APASL-positive patients without baseline A-TANGO ACLF: sHR 2.254 (1.886-2.723), p<0.001. These data identify A-TANGO-negative/APASL-positive patients as a high-risk pre-ACLF population **(Figure 4).**

**Figure 4:**
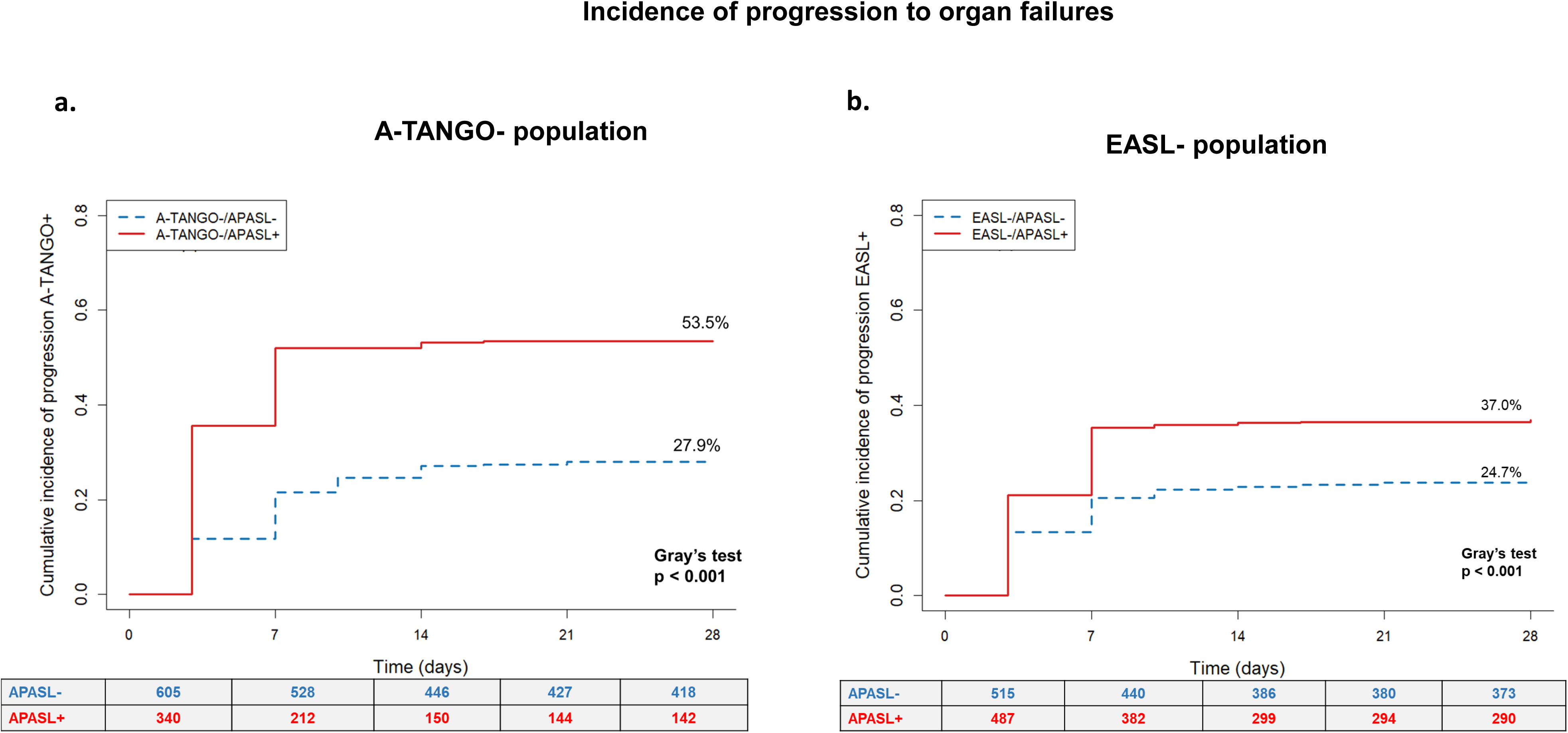
Cumulative incidence of progression to ACLF using competing-risk analysis. Cumulative incidence curves demonstrating progression to organ failure-defined ACLF over 28 days among baseline non-ACLF populations. (a) Progression to A-TANGO ACLF among A-TANGO-negative patients stratified by APASL status. (b) Progression to EASL-CLIF ACLF among EASL-negative patients stratified by APASL status. Fine-Gray competing-risk analysis was used, with death prior to ACLF onset treated as a competing event. Differences between groups were assessed using Gray’s test. Numbers at risk are shown below the curves. A two-sided p-value <0.05 was considered statistically significant.

#### Predictors of progression to A-TANGO ACLF

In univariable logistic regression among A-TANGO-negative patients, APASL positivity was strongly associated with progression to A-TANGO ACLF. Other variables associated with progression included ascites, higher A-TANGO OF score, bilirubin, INR, leukocyte count, lower sodium and albumin, and infection **(Table 2).**

**Table 2:** Predictors of progression to A-TANGO ACLF within A-TANGO-population at baseline.

| **Univariable analysis** |  |  |
| --- | --- | --- |
| **Variable** | **sHR (95% CI)** | **p-value** |
| Presence of APASL ACLF | 2.254 (1.886 – 2.723) | <0.001 |
| Presence of ascites | 1.412 (0.928 – 2.149) | 0.107 |
| A-TANGO OF | 1.056 (1.018 – 1.096) | 0.004 |
| Bilirubin | 1.022 (1.012 – 1.032) | <0.001 |
| INR | 1.126 (1.024 – 1.238) | 0.014 |
| Creatinine | 0.973 (0.875 – 1.082) | 0.613 |
| Sodium | 0.998 (0.995 – 1.000) | 0.049 |
| Albumin | 0.808 (0.681 - 0.959) | 0.015 |
| WBC | 1.019 (1.007 - 1.031) | 0.002 |
| Presence of Infection | 1.379 (1.018 – 1.715) | 0.004 |
| **Multivariable analysis (Models adjusted for age, gender and etiology of cirrhosis)** |  |  |
|  | **Adj. sHR** | **p-value** |
| **Model 1** |  |  |
| Presence of APASL ACLF | 2.306 (1.909 - 2.790) | <0.001 |
| A-TANGO OF score | 1.059 (1.015 – 1.100) | 0.008 |
| **Model 2** |  |  |
| Presence of APASL ACLF | 2.298 (1.903 – 2.780) | <0.001 |
| Presence of infection | 1.370 (1.099 – 1.710) | 0.005 |
| **Model 3** |  |  |
| Presence of Infections | 1.424 (1.142 – 1.770) | 0.002 |
| A-TANGO OF score | 1.061 (1.022 – 1.100) | 0.002 |
| **Model 4** |  |  |
| A-TANGO score | 1.040 (1.001 – 1.080) | 0.043 |
| WBC | 1.015 (1.003 - 1.030) | 0.015 |
| **Model 5** |  |  |
| Presence of APASL ACLF | 2.296 (1.901 – 2.770) | <0.001 |
| Presence of infection | 1.402 (1.124 – 1.750) | 0.003 |
| A-TANGO OF score | 1.065 (1.020 – 1.110) | 0.004 |
| **Model 6 (not adjusted for age, gender)** |  |  |
| A-TANGO OF score | 1.065 (1.020 – 1.111) | 0.001 |
| Presence of Infection | 1.422 (1.114 – 1.770) | 0.002 |
| MASLD as etiology of cirrhosis | 0.775 (0.520 – 1.160) | 0.210 |
| Subdistribution hazard ratios (SHRs) were estimated using Fine–Gray competing-risk regression models, with death prior to ACLF onset treated as a competing event.<br>Progression was defined as development of A-TANGO ACLF within 28 days among patients classified as A-TANGO– at baseline.<br>Multivariable models were adjusted for age, sex, and etiology of cirrhosis unless otherwise specified. Variables demonstrating collinearity with composite severity scores were not included simultaneously in adjusted models.<br>SHR, subdistribution hazard ratio; WBC, white blood cell count; OF, organ failure. |  |  |

In multivariable competing-risk analysis, APASL ACLF remained an independent predictor of progression to A-TANGO ACLF, with adjusted sHRs consistently around 2.30 across models. Progression was also independently associated with infection (sHR 1.402, 95% CI 1.124–1.750; p=0.003) and higher A-TANGO OF score (sHR 1.065, 95% CI 1.020–1.110; p=0.004) **(Table 2).** These findings suggest that APASL identifies a high-risk hepatic dysfunction phenotype, while infection and increasing organ-dysfunction burden further drive transition to A-TANGO ACLF.

#### Progression within ATANGO-/APASL-population

Among patients who were APASL-negative/A-TANGO-negative at baseline, follow-up data were available for 605 patients; 169 (27.9%) progressed to A-TANGO ACLF. Progressors were older and had more advanced decompensation, with higher rates of ascites (98.8% vs 92.7%), hepatic encephalopathy (90.5% vs 64.9%), infection (42.0% vs 14.2%), and tense ascites (48.5% vs 33.5%). Progression was accompanied by greater systemic and hepatic dysfunction, including higher WBC count, lower platelets, higher bilirubin, higher creatinine, lower sodium, and higher MELD, MELD-Na, A-TANGO OF, and AARC scores. Clinically, progression identified a very high-risk subgroup: 28-day mortality was 49.7% in progressors versus 4.1% in non-progressors, and 90-day mortality was 55.0% versus 6.7% **(Table S4)**.

A-TANGO OF score was the strongest and most consistent predictor of progression. On univariable analysis, progression was associated with A-TANGO OF score (sHR 1.121, 95% CI 1.070-1.175, p<0.001), bilirubin, creatinine, lower sodium, lower albumin, WBC count, and infection sHR 1.598, 95% CI 1.160 – 2.200, p=0.004). In multivariable models, A-TANGO OF score and infection remained independently associated with progression; in the final model, A-TANGO OF score had an adjusted sHR of 1.140, infection an adjusted sHR of 1.886 **(Table S5).** An A-TANGO OF threshold of ≥8 enriched for progression (52% vs. 19%), supporting its use to identify an additional pre-ACLF population suitable for prevention or enrichment studies.

### Reversal of A-TANGO ACLF

Among patients with A-TANGO ACLF at admission and evaluable follow-up, reversal to A-TANGO-negative status occurred in 238 patients (17.1%). Reversal was numerically less frequent among A-TANGO-positive/APASL-positive patients than among A-TANGO-positive/APASL-negative patients: 90 of 602 patients (15.0%) versus 148 of 786 patients (18.8%), respectively. Patients who achieved A-TANGO reversal had lower baseline systemic severity, including lower INR, creatinine, leukocyte count, MELD, MELD-Na, A-TANGO OF, and AARC scores **(Table S6).** They also had lower frequencies of cerebral, coagulation, circulatory, and respiratory failure. Liver failure prevalence was similar in reversal and non-reversal groups, suggesting that short-term reversibility was driven more by the burden of extrahepatic organ dysfunction than by hepatic injury alone. Mortality was substantially lower among patients who achieved reversal, with 28-day mortality of 9.2% compared with 62.5% among non-reversal patients. Competing-risk analysis, with death treated as a competing event, showed lower cumulative incidence of A-TANGO reversal among APASL-positive patients than APASL-negative patients 0.756 (0.586 – 0.975), p=0.031 **(Figure 5).**

**Figure 5:**
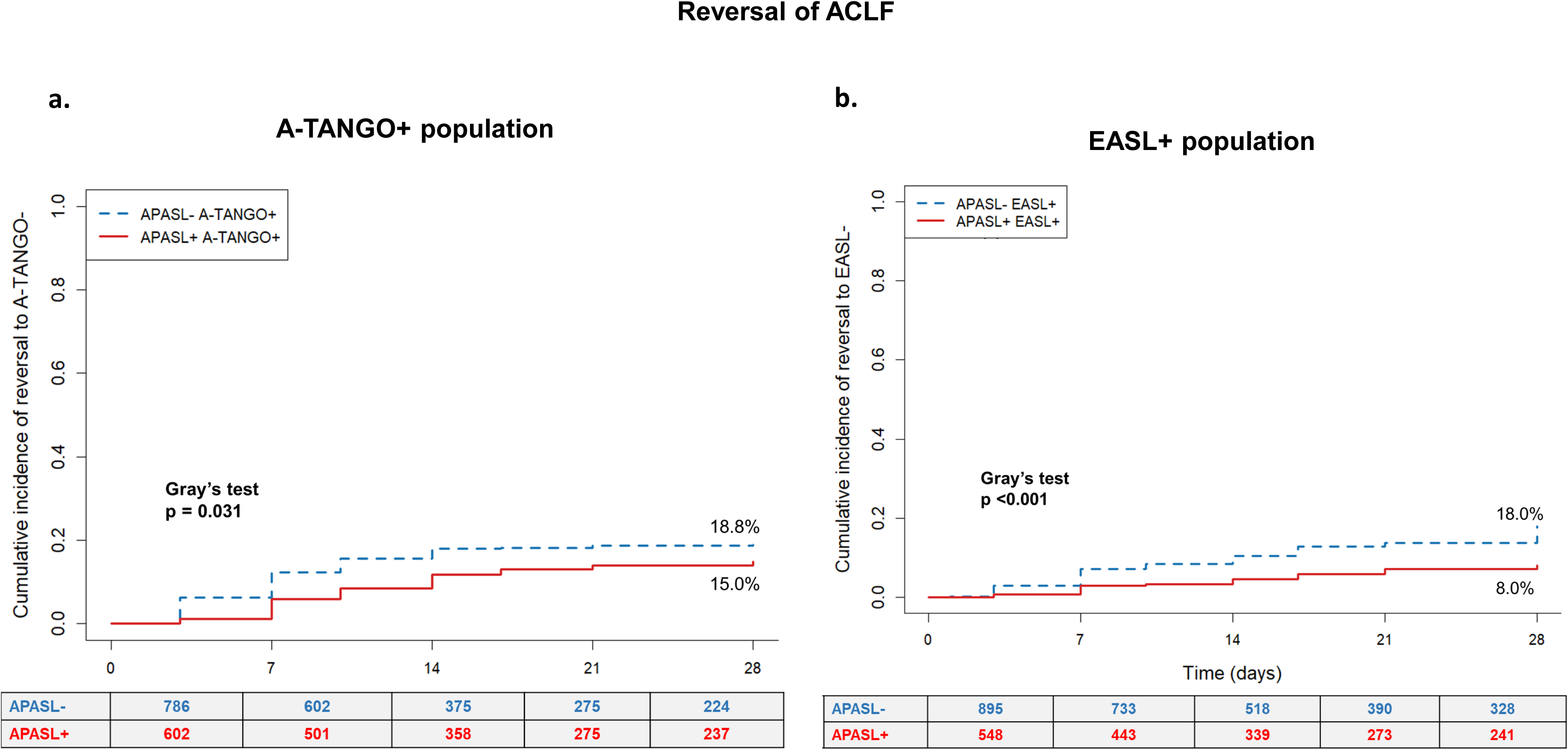
Cumulative incidence of ACLF reversal using competing-risk analysis. Cumulative incidence curves demonstrating reversal of organ failure-defined ACLF over 28 days among baseline ACLF populations. (a) Reversal of A-TANGO ACLF among A-TANGO+ patients stratified by APASL status. (b) Reversal of EASL-CLIF ACLF among EASL+ patients stratified by APASL status. Fine-Gray competing-risk analysis was used, with death prior to reversal treated as a competing event. Differences between groups were assessed using Gray’s test. Numbers at risk are shown below the curves. A two-sided p-value <0.05 was considered statistically significant.

#### Predictors of reversal in A-TANGO ACLF

On univariable competing-risk analysis, lower probability of reversal was associated with APASL ACLF (sHR 0.756, p=0.031), higher A-TANGO OF score (sHR 0.699, p<0.001), higher INR, creatinine, bilirubin, and the presence of cerebral, circulatory, respiratory, and coagulation failure. In adjusted models, A-TANGO OF score remained the most consistent predictor of reduced reversal (adj. sHR 0.701, p<0.001). Specific organ failures also strongly limited recovery, particularly circulatory failure (adj. sHR 0.445, p<0.001) and respiratory failure (adj. sHR 0.243, p<0.001) **(Table 3).** APASL ACLF was associated with lower reversal in models incorporating individual organ failures, suggesting that concomitant hepatic dysfunction further reduces short-term recovery once A-TANGO ACLF is established.

**Table 3:** Predictors of reversal (*A-TANGO criteria)* within *A-TANGO+* population at baseline.

|  | **Univariable analysis** |  |
| --- | --- | --- |
| **Variable** | **sHR (95% CI)** | **p-value** |
| Presence of APASL ACLF | 0.756 (0.586 – 0.975) | 0.031 |
| A-TANGO OF | 0.699 (0.651 - 0.750) | <0.001 |
| Bilirubin | 0.987 (0.976 – 0.999) | 0.032 |
| INR | 0.734 (0.633 - 0.852) | <0.001 |
| Creatinine | 0.885 (0.806 - 0.971) | 0.010 |
| WBC | 0.981 (0.962 – 1.001) | 0.062 |
| Platelet count | 1.002 (1.001 - 1.003) | 0.004 |
| Cerebral failure | 0.582 (0.403 - 0.839) | 0.004 |
| CVS failure | 0.455 (0.322 - 0.642) | <0.001 |
| Respiratory failure | 0.258 (0.144 - 0.461) | <0.001 |
| Coagulation failure | 0.572 (0.444 - 0.737) | <0.001 |
| **Multivariable analysis (Models adjusted for age, gender and etiology of cirrhosis)** |  |  |
|  | **Adj. sHR** | **p-value** |
| **Model 1** |  |  |
| Presence of APASL ACLF | 0.944 (0.731 - 1.218) | 0.656 |
| A-TANGO OF score | 0.701 (0.654 - 0.752) | <0.001 |
| **Model 2** |  |  |
| Presence of APASL ACLF | 0.731 (0.565 – 0.944) | 0.016 |
| Presence of CVS failure | 0.445 (0.314 - 0.631) | <0.001 |
| **Model 3** |  |  |
| A-TANGO OF score | 0.702 (0.653 - 0.754) | <0.001 |
| WBC | 0.995 (0.977 - 1.012) | 0.597 |
| **Model 4** |  |  |
| Presence of APASL ACLF | 0.766 (0.594 – 0.987) | 0.039 |
| WBC | 0.981 (0.962 – 1.001) | 0.061 |
| **Model 5** |  |  |
| Presence of APASL ACLF | 0.773 (0.600 – 0.997) | 0.048 |
| Creatinine | 0.890 (0.810 - 0.977) | 0.015 |
| **Model 6** |  |  |
| Presence of APASL ACLF | 0.731 (0.565 - 0.944) | 0.016 |
| CVS failure | 0.445 (0.314 – 0.631) | <0.001 |
| **Model 7** |  |  |
| Presence of APASL ACLF | 0.699 (0.543 - 0.900) | 0.006 |
| Respiratory failure | 0.243 (0.136 - 0.435) | <0.001 |
| Subdistribution hazard ratios (sHRs) were estimated using Fine–Gray competing-risk regression models, with death prior to reversal treated as a competing event.<br>Reversal was defined as resolution of A-TANGO ACLF within 28 days among patients classified as A-TANGO+ at baseline.<br>Multivariable models were adjusted for age, sex, and etiology of cirrhosis unless otherwise specified. Variables demonstrating collinearity with composite severity scores were not included simultaneously in adjusted models.<br>sHR, subdistribution hazard ratio; WBC, white blood cell count; OF, organ failure; CVS, cardiovascular. |  |  |

#### APASL-defined reversal

It was rare. Among APASL-positive patients with evaluable follow-up, APASL reversal occurred in 31 patients (4.0%). In the harmonized phenotype analysis, APASL reversal was observed in 15 of 156 A-TANGO-negative/APASL-positive patients (9.6%) and 16 of 612 A-TANGO-positive/APASL-positive patients (2.6%). Patients who reversed by APASL criteria had lower baseline bilirubin, lower A-TANGO OF and AARC scores, less liver failure, and lower mortality than those who did not reverse **(Table S7).** For APASL-defined reversal, lower bilirubin, lower systemic severity scores, higher sodium, and absence of liver failure were associated with greater likelihood of reversal in univariable analysis. Liver failure emerged as the most consistent negative predictor in adjusted analyses, consistent with the stringent biochemical requirements for APASL reversal **(Table 4).**

**Table 4:** Predictors of reversal (APASL criteria) within *APASL+* population at baseline.

|  | **Univariable analysis** |  |
| --- | --- | --- |
| **Variable** | **sHR (95% CI)** | **p-value** |
| A-TANGO OF score | 0.747 (0.620–0.899) | 0.002 |
| AARC score | 0.669 (0.536–0.835) | <0.001 |
| CLIF-C OF score | 0.767 (0.628–0.935) | 0.009 |
| Bilirubin | 0.894 (0.844–0.947) | <0.001 |
| Sodium | 1.075 (1.030–1.123) | 0.001 |
| Creatinine | 0.937 (0.645–1.362) | 0.734 |
| WBC | 0.870 (0.802–0.943) | <0.001 |
| Presence of HE | 0.262 (0.130–0.530) | <0.001 |
| Liver failure | 0.458 (0.290–0.723) | <0.001 |
| Coagulation failure | 0.549 (0.348–0.866) | 0.010 |
| **Multivariable analysis (Models adjusted for age, gender and etiology of cirrhosis)** |  |  |
|  | **Adj. sHR** | **p-value** |
| **Model 1** |  |  |
| CLIF-C OF score | 0.865 (0.680–1.101) | 0.238 |
| WBC | 0.877 (0.801–0.960) | 0.005 |
| **Model 2** |  |  |
| A-TANGO OF score | 0.823 (0.644–1.052) | 0.120 |
| WBC | 0.883 (0.805–0.970) | 0.009 |
| **Model 3** |  |  |
| AARC score | 0.668 (0.525–0.851) | 0.001 |
| WBC | 0.877 (0.788–0.976) | 0.016 |
| **Model 4** |  |  |
| Liver failure | 0.487 (0.238–0.997) | 0.049 |
| WBC | 0.878 (0.806–0.956) | 0.003 |
| **Model 5** |  |  |
| Coagulation failure | 0.484 (0.267–0.878) | 0.017 |
| WBC | 0.867 (0.792–0.948) | 0.002 |
| **Model 4** |  |  |
| Liver failure | 0.509 (0.243–1.064) | 0.073 |
| Coagulation failure | 0.504 (0.278–0.914) | 0.024 |
| WBC | 0.875 (0.799–0.957) | 0.004 |

Subdistribution hazard ratios (SHRs) were estimated using Fine–Gray competing-risk regression models, with death prior to reversal treated as a competing event.
APASL reversal was defined as resolution of APASL-defined hepatic dysfunction within 28 days, including bilirubin <5 mg/dL, INR <1.5, and resolution/control of ascites and hepatic encephalopathy among patients classified as APASL+ at baseline.
Multivariable models were adjusted for age, sex, and etiology of cirrhosis unless otherwise specified. Variables demonstrating collinearity with composite severity scores were not included simultaneously in adjusted models.
SHR, subdistribution hazard ratio; WBC, white blood cell count; OF, organ failure; HE, hepatic encephalopathy; AARC, APASL ACLF Research Consortium.

### Sensitivity analysis using EASL-CLIF criteria

Sensitivity analyses using EASL-CLIF criteria were directionally consistent with the primary A-TANGO analysis. Patients were reclassified into EASL-negative/APASL-negative, EASL-negative/APASL-positive, EASL-positive/APASL-negative, and EASL-positive/APASL-positive phenotypes. The distribution again showed substantial overlap between APASL-defined hepatic dysfunction and organ failure-based ACLF **(Table S8).** However, A-TANGO identified a slightly larger APASL high-risk organ failure population than EASL-CLIF.

Survival analyses showed clear prognostic separation across EASL/APASL phenotypes. Compared with EASL-negative/APASL-negative patients, mortality risk was higher in EASL-negative/APASL-positive patients and markedly higher in both EASL-positive groups **(Figure S1).** Among EASL-negative patients at baseline, APASL positivity was associated with higher 28-day progression to EASL-CLIF ACLF: 171 of 487 patients (35.1%) versus 123 of 515 patients (23.9%) in EASL-negative/APASL-negative patients **(Figure S2, Table S9-10).** Reversal by EASL criteria was less frequent among EASL-positive/APASL-positive patients than EASL-positive/APASL-negative patients: 44 of 548 patients (8.0%) versus 161 of 895 patients (18.0%). APASL-defined reversal remained infrequent in the EASL sensitivity analysis **(Table S11).**

Overall, the EASL-CLIF sensitivity analysis supported the consistency of the primary A-TANGO findings: APASL identifies a small but clinically important pre-ACLF phenotype when organ failure is absent, whereas outcomes after organ failure are driven predominantly by the burden and reversibility of systemic organ dysfunction.

## DISCUSSION

In this multicentre cohort of 4,024 patients hospitalised with acute decompensation of cirrhosis, we applied APASL and A-TANGO criteria simultaneously to test whether the two definitions describe competing or complementary populations. The data support a single coherent answer. APASL identifies a small but high-risk hepatic dysfunction phenotype that sits upstream of organ failure, A-TANGO defines the organ failure phase that dominates short-term mortality, and the two frameworks together describe a staged disease trajectory rather than two competing diagnoses. We summarise the findings under three themes, followed by their clinical and research implications.

APASL and A-TANGO criteria describe different clinical states that map onto a single mortality gradient. The A-TANGO-negative/APASL-positive phenotype was uncommon (8.7% of the cohort) but was clinically and biochemically distinct from acute decompensation without ACLF, with substantially higher bilirubin, higher INR, higher MELD-Na, and a clear predominance of hepatic precipitants such as alcohol-associated hepatitis and viral hepatitis. Despite this severity, 28-day mortality in A-TANGO-negative/APASL-positive patients were not statistically different from A-TANGO-negative/APASL-negative patients (15.1% vs 11.8%, p=0.128), although 90-day mortality was significantly higher (22.3% vs 14.4%, p=0.001). This temporal pattern indicates that APASL identifies hepatic injury before the mortality penalty of organ failure has accrued. Once A-TANGO organ failure was present, mortality rose sharply, to 45.4% at 28 days in A-TANGO-positive/APASL-positive patients and 56.0% in A-TANGO-positive/APASL-negative patients, with the highest hazard observed in the A-TANGO-positive/APASL-negative phenotype. Kaplan-Meier analysis showed clear and progressive separation across the four phenotypes. The interpretation is clear. Organ failure, defined by A-TANGO or EASL-CLIF, remains the dominant determinant of patient’s outcomes and short-term mortality, supported by literature(12–14). The presence of APASL-defined hepatic dysfunction on top of established organ failure does not add further prognostic information for death. APASL therefore informs a state before organ failures, A-TANGO defines the phase of organ failure, and these two frameworks are stages of one disease spectrum rather than alternative diagnoses.

Our interpretation is supported by observation that APASL positivity identifies the patients who progress to organ failure-defined ACLF. Among A-TANGO-negative patients with evaluable follow-up, 53.5% of APASL-positive patients progressed to A-TANGO ACLF within 28 days, compared with 27.9% of APASL-negative patients. In competing-risk analysis with death treated as a competing event, APASL positivity remained an independent driver of progression, with an adjusted subdistribution hazard ratio of approximately 2.30 across multivariable models adjusted for age, sex and aetiology of cirrhosis. Infection (adjusted sHR 1.40) and baseline A-TANGO OF score (adjusted sHR 1.07 per point) were independent co-drivers, but APASL positivity was consistently the strongest predictor of progression. These findings align closely with prior studies demonstrating that baseline liver dysfunction, circulatory derangements, and systemic inflammation are major determinants of ACLF development and early progression(15, 16). The strong contribution of infection and leukocytosis in our cohort further supports the concept of a “golden window” preceding overt organ failure, during which systemic inflammation and sepsis-related immune dysregulation drive transition from hepatic decompensation to established ACLF(17). The EASL-CLIF sensitivity analysis reproduced the same pattern, with 35.1% progression in EASL-negative/APASL-positive patients versus 23.9% in EASL-negative/APASL-negative patients, indicating that the progression is linked to presence of APASL positivity itself and not of any specific organ failure definition.

A second and complementary observation came from the A-TANGO-negative/APASL-negative subgroup. Even in patients without APASL-defined hepatic dysfunction, an A-TANGO OF score of 8 or higher enriched for subsequent progression (52% vs 19%), with A-TANGO OF score and infection emerging as the strongest predictors in multivariable competing-risk models. Two distinct pre-ACLF populations are therefore identifiable at admission. The first is the A-TANGO-negative/APASL-positive patient, in whom hepatic injury dominates and progression is driven by the trajectory of liver dysfunction. The second is the A-TANGO-negative/APASL-negative patient with an A-TANGO OF score of 8 or higher, in whom systemic dysfunction is starting to declare itself without yet meeting organ failure criteria. Both groups are clinically tractable targets for prevention and for trial enrichment. These findings are consistent with prior observations that progression to ACLF may already be detectable within apparently uncomplicated acute decompensation through the presence of early systemic dysfunction and heightened inflammatory burden(18). They further support the concept that ACLF is initiated by diverse precipitating events which converge on common inflammatory and haemodynamic pathways leading to organ failure(5, 19, 20), and with the role of infection as a key amplifier of systemic inflammation in cirrhosis(21).

Reversal mirrors progression and reinforces the staged interpretation. A-TANGO ACLF reversed in 17.1% of evaluable patients overall, and the cumulative incidence of reversal was lower in APASL-positive patients than in APASL-negative patients (sHR 0.756, 95% CI 0.586 to 0.975, p=0.031). In adjusted competing-risk models, APASL positivity reduced the probability of A-TANGO reversal in every model that incorporated individual organ failures, although the global A-TANGO OF score remained the single most consistent determinant of recovery (adjusted sHR 0.701 per point). Specific organ failures had a dominant effect, particularly circulatory failure (adjusted sHR 0.445) and respiratory failure (adjusted sHR 0.243), highlighting the dominant influence of extrahepatic organ dysfunction once ACLF is established. These observations are concordant with prior longitudinal ACLF studies showing that reversal or improvement occurs in only a subset of patients. In the CANONIC cohort, ACLF resolved in 42.5% of patients within 28-days and was linked to markedly better survival, while persistence of organ failure was associated with high short-term mortality(22). CAP-ACLF from APASL ACLF research consortium also demonstrated that reversal of ACLF within the first 7 days was strongly associated with improved survival, whereas failure of reversal or progression of ACLF grade carried markedly poor outcomes, emphasizing the prognostic importance of early recovery from organ failure(12). The EASL-CLIF sensitivity analysis showed the same direction of effect, with EASL ACLF reversal in 18.0% of EASL-positive/APASL-negative patients but only 8.0% of EASL-positive/APASL-positive patients.

APASL-defined reversal was rare, occurring in only 4.0% of APASL-positive patients overall, with a clear gradient by baseline organ failure status: 9.6% in A-TANGO-negative/APASL-positive patients and only 2.6% in A-TANGO-positive/APASL-positive patients. Lower bilirubin, lower A-TANGO OF score, higher sodium, and absence of liver failure were associated with APASL reversal in univariable analysis, with liver failure and coagulation failure persisting as negative predictors in adjusted models. This finding was consistent with short-term reversal rates of 2.3% in APASL-ACLF from the studies emanating from AARC consortium (12). The stringency of APASL reversal, which requires reduction bilirubin and INR to less than 5 and 1.5, as well as resolution or stabilisation of ascites and encephalopathy, makes it inevitably less attainable than reversal of organ failure alone; especially in the short-term. On the other hand, long-term follow-up studies of APASL-ACLF due to hepatitis B virus infection demonstrate that hepatic recovery occur gradually over prolonged follow-up of 5 years among survivors, rather than during the early acute phase captured in short-term analyses(23). Taken together, the two reversal analyses show that established hepatic dysfunction reduces the probability of organ failure reversal, that established organ failure reduces the probability of hepatic dysfunction reversal, and that the two definitions are reciprocally informative for short-term recovery.

These findings argue for a staged and harmonised approach to ACLF rather than a choice between definitions. Within this framework, A-TANGO-negative/APASL-negative patients with low organ-dysfunction burden represent relatively stable acute decompensation; A-TANGO-negative/APASL-positive patients represent a hepatic dysfunction-dominant, progression-enriched pre-ACLF phase; A-TANGO-negative/APASL-negative patients with an A-TANGO OF score of 8 or higher represent a systemic vulnerability pre-ACLF phenotype; and A-TANGO-positive patients, with or without APASL positivity, represent established organ failure-defined ACLF. Two clinical priorities follow directly. First, A-TANGO-negative/APASL-positive patients and A-TANGO-negative/APASL-negative patients with an A-TANGO OF score of 8 or higher are the populations in whom infection surveillance, early management of precipitating events, nephrotoxin avoidance, and timely escalation of care are most likely to alter the trajectory. Second, patients with established A-TANGO ACLF, particularly those who are also APASL-positive, should be triaged for organ support and early transplant evaluation, recognising that spontaneous reversal is uncommon and is further reduced by concomitant hepatic dysfunction. From a research perspective, these two pre-ACLF populations are natural enrichment targets for prevention trials, and the harmonised four-phenotype framework provides a coherent basis for aligning therapeutic strategy with disease phase.

The strengths of this analysis include the large cohort size, the simultaneous application of APASL, A-TANGO and EASL-CLIF criteria, longitudinal characterisation of progression and reversal, and the use of competing-risk methods with death treated as a competing event. The consistency of findings across A-TANGO and EASL-CLIF sensitivity analyses supports robustness. Several limitations should be acknowledged. This cohort was derived from tertiary care centres in India and included a predominance of alcohol-associated liver disease; therefore, replication of these findings in cohorts with other underlying liver disease aetiologies is needed. Longitudinal data on progression and reversal were available only in a prospectively recruited and carefully characterised subgroup. However, because clinical outcomes and mortality rates in this subgroup were similar to those observed among patients without complete progression–reversal data, we believe this limitation is unlikely to have materially altered the interpretation of the findings. Nevertheless, external validation in geographically diverse and aetiologically distinct cohorts is required before the proposed staged framework can be considered definitive.

In conclusion, APASL-defined ACLF does not compete with A-TANGO or EASL-CLIF; it occupies an upstream position on the same disease trajectory. The A-TANGO-negative/APASL-positive phenotype, although uncommon, identifies a high-risk pre-ACLF population in whom more than half of patients progress to A-TANGO ACLF within 28 days. An A-TANGO OF score of 8 or higher within the A-TANGO-negative/APASL-negative subgroup identifies a second, complementary pre-ACLF population. Once organ failure is established, short-term mortality is dominated by A-TANGO criteria, and APASL positivity at baseline independently reduces the probability of organ failure reversal. These data support an integrated and staged ACLF framework, with direct implications for early risk stratification, surveillance, and the design of prevention-focused clinical trials.

## Data Availability

All data produced in the present study are available upon reasonable request to the authors

## Notes

### Competing Interest Statement

The authors have declared no competing interest.

### Author Declarations

Institutional Ethics Committee at PGIMER Chandigarh has approved for this work (PGI/IEC-08/2021-2062).

## REFERENCES

1. Sarin SK, Choudhury A, Kumar A, Mahmud N, Lee GH, Ning Q, et al. Acute-on-chronic liver failure: pathophysiological mechanisms and clinical management. Nat Rev Gastroenterol Hepatol. 2026;23(5):411–31.

2. Moreau R, Jalan R, Gines P, Pavesi M, Angeli P, Cordoba J, et al. Acute-on-chronic liver failure is a distinct syndrome that develops in patients with acute decompensation of cirrhosis. Gastroenterology. 2013;144(7):1426–37, 37.e1-9.

3. O’Leary JG, Reddy KR, Garcia-Tsao G, Biggins SW, Wong F, Fallon MB, et al. NACSELD acute-on-chronic liver failure (NACSELD-ACLF) score predicts 30-day survival in hospitalized patients with cirrhosis. Hepatology. 2018;67(6):2367–74.

4. Sarin SK, Kedarisetty CK, Abbas Z, Amarapurkar D, Bihari C, Chan AC, et al. Acute-on-chronic liver failure: consensus recommendations of the Asian Pacific Association for the Study of the Liver (APASL) 2014. Hepatol Int. 2014;8(4):453–71.

5. Choudhury A, Kulkarni AV, Arora V, Soin AS, Dokmeci AK, Chowdhury A, et al. Acute-on-chronic liver failure (ACLF): the ‘Kyoto Consensus’-steps from Asia. Hepatol Int. 2025;19(1):1–69.

6. Engelmann C, Verma N, Qi T, Catharina Kerbert AJ, Pohl J, Morgan C, et al. Development and validation of the A-TANGO organ failure score for acute-on-chronic liver failure in global cohorts. J Hepatol. 2026.

7. Bajaj JS, Shawcross DL, Choudhury A, Karvellas CJ, O’Leary JG, Trebicka J, et al. Defining Organ Failures in Patients With Cirrhosis: Consensus Statements. Gastroenterology. 2025;169(5):1043–62.

8. Verma N, Qi T, Garg P, Valsan A, Nair G, Pohl J, et al. Consensus versus Outcome-based Definitions of Acute-on-chronic Liver Failure: Implications for Risk Stratification and Case Identification. Journal of Clinical and Translational Hepatology. 2026(000).

9. Meiqian Hu JL, Nipun Verma, Pratibha Garg, Sunil Taneja, Juan Antonio Carbonell-Asins, María Pilar Ballester, Tingting Qi, Sina Jameie-Oskooei, Qun Cai, Xi Liang, Jiaqi Li, Tianzhou Wu, Jiang Li, Peng Li, Qian Zhou, Jiaojiao Xin, Dongyan Shi, Jing Jiang, Wei Qiang, Changze Hong, Xin Chen, Bing Zhu, Tingting Feng, Jianming Zheng, Yuxian Huang, Feng Ye, Bingliang Lin, Jinjun Chen, Rajeshwar P Mookerjee, Yan Huang, Shaoli You, Cornelius Engelmann, Yu Chen, Ajay Duseja, Jun Li, Rajiv Jalan. A Comparison of Diagnostic Models and Prognostic Scores of ACLF: Towards Global Harmonization. 2026.

10. Jalan R, Qi T, Engelmann C, Chen J, Li H, Li J, et al. The Illusion of Harmonisation in ACLF. J Hepatol. 2026.

11. Lee BP, Cullaro G, Vosooghi A, Yao F, Panchal S, Goldberg DS, et al. Discordance in categorization of acute-on-chronic liver failure in the United Network for Organ Sharing database. J Hepatol. 2022;76(5):1122–6.

12. Verma N, Dhiman RK, Singh V, Duseja A, Taneja S, Choudhury A, et al. Comparative accuracy of prognostic models for short-term mortality in acute-on-chronic liver failure patients: CAP-ACLF. Hepatol Int. 2021;15(3):753–65.

13. Jalan R, Saliba F, Pavesi M, Amoros A, Moreau R, Ginès P, et al. Development and validation of a prognostic score to predict mortality in patients with acute-on-chronic liver failure. J Hepatol. 2014;61(5):1038–47.

14. Choudhury A, Jindal A, Maiwall R, Sharma MK, Sharma BC, Pamecha V, et al. Liver failure determines the outcome in patients of acute-on-chronic liver failure (ACLF): comparison of APASL ACLF research consortium (AARC) and CLIF-SOFA models. Hepatol Int. 2017;11(5):461–71.

15. Piano S, Tonon M, Vettore E, Stanco M, Pilutti C, Romano A, et al. Incidence, predictors and outcomes of acute-on-chronic liver failure in outpatients with cirrhosis. J Hepatol. 2017;67(6):1177–84.

16. Verma N, Roy A, Valsan A, Garg P, Ralmilay S, Girish V, et al. Liver Dysfunction and Systemic Inflammation Drive Organ Failures in Acute Decompensation of Cirrhosis: A Multicentric Study. Am J Gastroenterol. 2025;120(1):182–93.

17. Choudhury A, Kumar M, Sharma BC, Maiwall R, Pamecha V, Moreau R, et al. Systemic inflammatory response syndrome in acute-on-chronic liver failure: Relevance of ‘golden window’: A prospective study. J Gastroenterol Hepatol. 2017;32(12):1989–97.

18. Trebicka J, Fernandez J, Papp M, Caraceni P, Laleman W, Gambino C, et al. The PREDICT study uncovers three clinical courses of acutely decompensated cirrhosis that have distinct pathophysiology. J Hepatol. 2020;73(4):842–54.

19. Arroyo V, Angeli P, Moreau R, Jalan R, Clària J, Trebicka J, et al. The systemic inflammation hypothesis: Towards a new paradigm of acute decompensation and multiorgan failure in cirrhosis. J Hepatol. 2021;74(3):670–85.

20. Clària J, Stauber RE, Coenraad MJ, Moreau R, Jalan R, Pavesi M, et al. Systemic inflammation in decompensated cirrhosis: Characterization and role in acute-on-chronic liver failure. Hepatology. 2016;64(4):1249–64.

21. Cullaro G, Sharma R, Trebicka J, Cárdenas A, Verna EC. Precipitants of Acute-on-Chronic Liver Failure: An Opportunity for Preventative Measures to Improve Outcomes. Liver Transpl. 2020;26(2):283–93.

22. Gustot T, Fernandez J, Garcia E, Morando F, Caraceni P, Alessandria C, et al. Clinical Course of acute-on-chronic liver failure syndrome and effects on prognosis. Hepatology. 2015;62(1):243–52.

23. Wang H, Tong J, Xu X, Chen J, Mu X, Zhai X, et al. Reversibility of acute-on-chronic liver failure syndrome in hepatitis B virus-infected patients with and without prior decompensation. J Viral Hepat. 2022;29(10):890–8.

