## Supplementary tables for "Harmonising APASL and A-TANGO criteria for acute-on-chronic liver failure: identification of complementary high-risk pre-ACLF populations": Supplementary tables.docx

| *Table 1: Baseline characteristics of cohort (N=4024)* | |
| --- | --- |
| Characteristic |  |
| Age, median (IQR) | 45.0 (38.0 - 54.0) |
| Female sex, n/N (%) | 440 (10.9) |
| Etiology of cirrhosis (n, %) |  |
| Alcohol | 2873 (71.4) |
| MASLD | 308 (7.7) |
| Hepatitis B/C | 363 (9.0) |
| Alcohol+ CMRF | 69 (1.7) |
| AIH | 191 (4.7) |
| Cryptogenic | 121 (3.0) |
| Alcohol+ Hepatitis B/C | 86 (2.1) |
| Others | 13 (0.3) |
| Precipitating events (n, %) |  |
| AH | 1119 (27.8) |
| Infections | 676 (16.8) |
| AH+ others | 1031 (25.6) |
| DILI | 226 (5.6) |
| Viral hepatitis | 250 (6.2) |
| AIH | 75 (1.9) |
| UGIB/LGIB | 475 (11.8) |
| Others | 172 (4.3) |
| Type of acute insult |  |
| Hepatic | 2701 (67.1) |
| Extra-hepatic | 1323 (32.9) |
| More than acute insult | 1435 (35.7) |
| Prior decompensation | 586 (20.1) |
| Decompensations |  |
| Prevalence of ascites | 3695 (91.8) |
| Ascites grade |  |
| No Ascites | 136 (3.7) |
| Mild to moderate | 2356 (64.9) |
| Tense ascites | 1136 (31.3) |
| Prevalence of HE | 2524 (62.7) |
| HE grades |  |
| No HE | 1181 (32.6) |
| Grade I-II | 2001 (55.2) |
| Grade III-IV | 446 (12.3) |
| Infections | 1315 (36.2) |
| Lab investigations |  |
| Haemoglobin (gm/dL) | 9.4 (7.8 - 11.1) |
| WBCC (x10^9^/L) | 10.1 (6.6 - 15.8) |
| Platelet count (x10^9^/L) | 99.0 (65.0 - 151.0) |
| INR | 1.9 (1.5 - 2.5) |
| AST (U/L) | 114.0 (71.0 - 184.9) |
| ALT (U/L) | 48.0 (30.0 - 89.0) |
| Bilirubin (mg/dL) | 12.6 (5.2 - 22.3) |
| Creatinine (mg/dL) | 1.1 (0.8 - 2.0) |
| Sodium (mmol/L) | 132.0 (128.0 - 136.0) |
| MAP (mmHg) | 83.7 (76.7 - 92.0) |
| Disease severity scores |  |
| MELD score | 25.6 (19.0 - 32.0) |
| MELD-Na score | 29.4 (24.3 - 34.5) |
| CLIF-C OF score | 9.0 (8.0 - 11.0) |
| ATANGO- OF score | 10.0 (8.0 - 12.0) |
| AARC score | 10.0 (8.0 - 11.0) |
| INR: international normalized ration, AST: Aspartate Aminotransferase, ALT: Aspartate Aminotransferase, MAP: Mean arterial pressure, MELD: Model of end-stage liver disease, MELD-Na: MELD-sodium  Categorical variables are presented as n (%), and continuous variables as median (interquartile range, IQR). Categorical variables were compared using the Chi-square test, and continuous variables using the Mann–Whitney U test. A two-sided p-value <0.05 was considered statistically significant. | |

| Table 2: Univariable Cox regression analysis for 28-day mortality across ACLF phenotypes defined as per A-TANGO criteria | | | |
| --- | --- | --- | --- |
| Phenotype | HR | 95% CI | p-value |
| A-TANGO-/APASL- | Reference | - | - |
| A-TANGO-/APASL+ | 1.34 | 0.96 – 1.85 | 0.082 |
| A-TANGO+/APASL- | 6.50 | 5.31 – 7.95 | <0.001 |
| A-TANGO+/APASL+ | 4.80 | 3.94 – 5.85 | <0.001 |

| *Table 3: Baseline characteristics of A-TANGO^-^ cohort stratified by disease progression over follow up* | | | |
| --- | --- | --- | --- |
| Characteristic | No progression  (n=594, 62.9%) | Progression to A-TANGO+  (n= 351, 37.1%) | P-value |
| Age, median (IQR) | 46.0 (40.0 - 53.0) | 46.0 (41.0 - 55.5) | 0.244 |
| Female sex, n/N (%) | 51 (8.6) | 42 (12.0) | 0.116 |
| Presence of APSAL-ACLF | 158 (26.6) | 182 (51.9) | <0.001 |
| Etiology of cirrhosis (n, %) |  |  |  |
| Alcohol | 476 (80.1) | 248 (70.7) | <0.001 |
| MASLD | 25 (4.2) | 38 (10.8) |  |
| Hepatitis B/C | 21 (3.5) | 38 (10.8) |  |
| Alcohol+ CMRF | 15 (2.5) | 2 (0.6) |  |
| AIH | 26 (4.4) | 12 (3.4) |  |
| Cryptogenic | 15 (2.5) | 8 (2.3) |  |
| Alcohol+ Hepatitis B/C | 13 (2.2) | 4 (1.1) |  |
| Others | 3 (0.5) | 1 (0.3) |  |
| Precipitating events (n, %) |  |  |  |
| AH | 158 (26.6) | 109 (31.1) | <0.001 |
| Infections | 105 (17.7) | 73 (20.8) |  |
| AH+ others | 222 (37.4) | 87 (24.8) |  |
| DILI | 33 (5.6) | 24 (6.8) |  |
| Viral hepatitis | 16 (2.7) | 35 (10.0) |  |
| AIH | 16 (2.7) | 4 (1.1) |  |
| UGIB/LGIB | 21 (3.5) | 5 (1.4) |  |
| Others | 23 (3.9) | 14 (4.0) |  |
| Type of acute insult |  |  |  |
| Hepatic | 445 (74.9) | 259 (73.8) | 0.759 |
| Extra-hepatic | 149 (25.1) | 92 (26.2) |  |
| More than acute insult | 245 (41.2) | 101 (28.8) | <0.001 |
| Prior decompensation | 23 (12.7) | 19 (9.5) | 0.394 |
| Decompensations |  |  |  |
| Prevalence of Ascites | 550 (92.6) | 339 (96.6) | 0.018 |
| Ascites grade |  |  |  |
| No Ascites | 44 (7.4) | 12 (3.4) | 0.038 |
| Mild to moderate | 363 (61.1) | 229 (65.2) |  |
| Tense ascites | 187 (31.5) | 110 (31.3) |  |
| Prevalence of HE | 374 (63.0) | 246 (70.1) | 0.031 |
| HE grades |  |  |  |
| No HE | 220 (37.0) | 105 (29.9) | <0.001 |
| Grade I | 324 (54.5) | 138 (39.3) |  |
| Grade II | 45 (7.6) | 106 (30.2) |  |
| Grade III | 5 (0.8) | 2 (0.6) |  |
| Grade IV | 0 (0.0) | 0 (0.0) |  |
| Infections | 90 (15.2) | 122 (34.8) | <0.001 |
| Lab investigations |  |  |  |
| Haemoglobin (gm/dL) | 9.8 (8.0 - 11.3) | 10.2 (8.2 - 11.7) | 0.112 |
| WBCC (x10^9^/L) | 7.7 (5.6 - 10.7) | 9.0 (6.1 - 13.0) | <0.001 |
| Platelet count (x10^9^/L) | 101.0 (66.0 - 150.0) | 96.0 (55.0 - 156.5) | 0.414 |
| INR | 1.4 (0.9 - 1.7) | 1.6 (1.2 - 1.9) | <0.001 |
| AST (U/L) | 87.0 (55.2 - 144.6) | 109.0 (69.2 - 179.0) | <0.001 |
| ALT (U/L) | 43.0 (27.0 - 70.3) | 45.4 (29.0 - 90.8) | 0.010 |
| Bilirubin (mg/dL) | 6.1 (3.4 - 10.3) | 9.7 (5.6 - 15.1) | <0.001 |
| Creatinine (mg/dL) | 0.8 (0.7 - 1.1) | 0.9 (0.7 - 1.2) | 0.018 |
| Sodium (mmol/L) | 134.0 (130.3 - 137.0) | 132.5 (128.0 - 136.0) | <0.001 |
| MAP (mmHg) | 87.0 (80.7 - 92.3) | 85.3 (78.0 - 92.0) | 0.112 |
| Disease severity scores |  |  |  |
| MELD score | 17.2 (13.3 - 21.0) | 21.0 (17.5 - 23.3) | <0.001 |
| MELD-Na score | 20.3 (16.2 - 24.1) | 24.4 (20.9 - 27.2) | <0.001 |
| ATANGO- OF score | 7.0 (6.0 - 8.0) | 8.0 (7.0 - 8.0) | <0.001 |
| AARC score | 8.0 (7.0 - 8.0) | 8.0 (7.0 - 9.0) | <0.001 |
| Organ failures as per ATAGO criteria |  |  |  |
| Cerebral failure | 5 (0.8) | 2 (0.6) | 0.937 |
| Coagulation failure | - | - | - |
| Liver failure | - | - | - |
| Kidney failure | - | - | - |
| Circulatory failure | 5 (0.8) | 7 (2.0) | 0.219 |
| Respiratory failure | - | - | - |
| 28-day mortality | 34 (5.7) | 119 (33.9) | <0.001 |
| 90-day mortality | 51 (8.6) | 146 (41.6) | <0.001 |
| INR: international normalized ration, AST: Aspartate Aminotransferase, ALT: Aspartate Aminotransferase, MAP: Mean arterial pressure, MELD: Model of end-stage liver disease, MELD-Na: MELD-sodium  Categorical variables are presented as n (%), and continuous variables as median (interquartile range, IQR). Categorical variables were compared using the Chi-square test, and continuous variables using the Mann–Whitney U test. A two-sided p-value <0.05 was considered statistically significant.  Complete follow up data till 28-days/death was available for 945 patients out of 1309 A-TANGO- patients at baseline | | | |

| *Table 4: Baseline characteristics of APASL-/A-TANGO- cohort stratified by disease progression over follow up* | | | |
| --- | --- | --- | --- |
| Characteristic | No progression  (n=436, 72.1%) | Progression to A-TANGO+  (n= 169, 27.9%) | p-value |
| Age, median (IQR) | 46.0 (40.0 - 54.0) | 49.0 (42.0 - 57.0) | 0.019 |
| Female sex, n/N (%) | 35 (8.0) | 21 (12.4) | 0.129 |
| Etiology of cirrhosis (n, %) |  |  |  |
| Alcohol | 348 (79.8) | 131 (77.5) | 0.001 |
| MASLD | 24 (5.5) | 27 (16.0) |  |
| Hepatitis B/C | 11 (2.5) | 4 (2.4) |  |
| Alcohol+ CMRF | 14 (3.2) | 0 (0.0) |  |
| AIH | 21 (4.8) | 4 (2.4) |  |
| Cryptogenic | 9 (2.1) | 1 (0.6) |  |
| Alcohol+ Hepatitis B/C | 6 (1.4) | 1 (0.6) |  |
| Others | 3 (0.7) | 1 (0.6) |  |
| Precipitating events (n, %) |  |  |  |
| AH | 83 (19.0) | 27 (16.0) | 0.187 |
| Infections | 90 (20.6) | 53 (31.4) |  |
| AH+ others | 178 (40.8) | 64 (37.9) |  |
| DILI | 26 (6.0) | 10 (5.9) |  |
| Viral hepatitis | 6 (1.4) | 3 (1.8) |  |
| AIH | 14 (3.2) | 4 (2.4) |  |
| UGIB/LGIB | 19 (4.4) | 4 (2.4) |  |
| Others | 20 (4.6) | 4 (2.4) |  |
| Type of acute insult |  |  |  |
| Hepatic | 307 (70.4) | 108 (63.9) | 0.147 |
| Extra-hepatic | 129 (29.6) | 61 (36.1) |  |
| More than acute insult | 198 (45.4) | 68 (40.2) | 0.289 |
| Prior decompensation | - | - | - |
| Decompensations |  |  |  |
| Prevalence of Ascites | 404 (92.7) | 167 (98.8) | 0.006 |
| Ascites grade |  |  |  |
| No Ascites | 32 (7.3) | 2 (1.2) | <0.001 |
| Mild to moderate | 258 (59.2) | 85 (50.3) |  |
| Tense ascites | 146 (33.5) | 82 (48.5) |  |
| Prevalence of HE | 283 (64.9) | 153 (90.5) | <0.001 |
| HE grades |  |  |  |
| No HE | 153 (35.1) | 16 (9.5) | <0.001 |
| Grade I | 243 (55.7) | 71 (42.0) |  |
| Grade II | 35 (8.0) | 80 (47.3) |  |
| Grade III | 5 (1.1) | 2 (1.2) |  |
| Infections | 62 (14.2) | 71 (42.0) | <0.001 |
| Lab investigations |  |  |  |
| Haemoglobin (gm/dL) | 9.8 (7.9 - 11.1) | 10.1 (7.8 - 11.3) | 0.789 |
| WBCC (x10^9^/L) | 7.6 (5.3 - 10.3) | 9.4 (6.1 - 13.6) | <0.001 |
| Platelet count (x10^9^/L) | 101.0 (64.8 - 149.0) | 78.0 (48.5 - 133.0) | 0.004 |
| INR | 1.1 (0.8 - 1.5) | 1.2 (0.8 - 1.5) | 0.090 |
| AST (U/L) | 78.2 (51.2 - 123.0) | 93.5 (58.0 - 164.0) | 0.009 |
| ALT (U/L) | 41.1 (25.7 - 64.0) | 40.9 (24.5 - 75.6) | 0.653 |
| Bilirubin (mg/dL) | 4.8 (2.7 - 9.2) | 8.0 (4.2 - 14.7) | <0.001 |
| Creatinine (mg/dL) | 0.8 (0.7 - 1.1) | 0.9 (0.8 - 1.2) | 0.010 |
| Sodium (mmol/L) | 134.2 (131.0 - 137.0) | 132.5 (126.0 - 137.0) | 0.001 |
| MAP (mmHg) | 87.0 (81.7 - 92.3) | 85.7 (78.0 - 91.3) | 0.142 |
| Disease severity scores |  |  |  |
| MELD score | 15.4 (12.4 - 17.9) | 17.3 (15.2 - 20.6) | <0.001 |
| MELD-Na score | 18.2 (14.4 - 21.9) | 22.0 (18.2 - 25.6) | <0.001 |
| A-TANGO OF score | 7.0 (6.0 - 7.0) | 8.0 (7.0 - 8.0) | <0.001 |
| AARC score | 7.0 (7.0 - 8.0) | 8.0 (7.0 - 9.0) | <0.001 |
| Organ failures as per A-TANGO criteria |  |  |  |
| Cerebral failure | 5 (1.1) | 2 (1.2) | 1.000 |
| Coagulation failure | - | - | - |
| Liver failure | - | - | - |
| Kidney failure | - | - | - |
| Circulatory failure | 4 (0.9) | 7 (4.1) | 0.020 |
| Respiratory failure | - | - | - |
| 28-day mortality | 18 (4.1) | 84 (49.7) | <0.001 |
| 90-day mortality | 29 (6.7) | 93 (55.0) | <0.001 |
| INR: international normalized ration, AST: Aspartate Aminotransferase, ALT: Aspartate Aminotransferase, MAP: Mean arterial pressure, MELD: Model of end-stage liver disease, MELD-Na: MELD-sodium  Categorical variables are presented as n (%), and continuous variables as median (interquartile range, IQR). Categorical variables were compared using the Chi-square test, and continuous variables using the Mann–Whitney U test. A two-sided p-value <0.05 was considered statistically significant.  Follow up data was available for 605 patients out of 959 APASL-/A-TANGO- patients at baseline | | | |

| *Table 5: Predictors of progression to A-TANGO ACLF within APASL-/A-TANGO- population at baseline* | | |
| --- | --- | --- |
|  | Univariable analysis | |
| Variable | sHR (95% CI) | p-value |
| A-TANGO OF | 1.121 (1.070 –1.175) | <0.001 |
| Bilirubin | 1.027 (1.012 –1.041) | <0.001 |
| Creatinine | 1.120 (0.998 –1.258) | 0.054 |
| Sodium | 0.995 (0.991 – 0.999) | 0.006 |
| Albumin | 0.850 (0.660 – 1.096) | 0.211 |
| WBC | 1.036 (1.020 – 1.052) | <0.001 |
| Presence of Infection | 1.598 (1.160 – 2.200) | 0.004 |
| Multivariable analysis (Models adjusted for age, gender and etiology of cirrhosis) | | |
|  | Adj. sHR | p-value |
| Model 1 | | |
| Presence of Infections | 1.886 (1.358 – 2.620) | <0.001 |
| A-TANGO OF score | 1.140 (1.086 – 1.197) | <0.001 |
| Model 2 | | |
| A-TANGO score | 1.098 (1.046 – 1.154) | <0.001 |
| WBC | 1.021 (1.002 – 1.040) | 0.029 |
| Model 3 | | |
| A-TANGO score | 1.132 (1.081 – 1.186) | <0.001 |
| Sodium | 0.995 (0.991 – 0.999) | 0.013 |
| Model 6 (not adjusted for age, gender) | | |
| A-TANGO OF score | 1.111 (1.056 – 1.170) | <0.001 |
| WBC | 1.021 (1.003 – 1.040) | 0.022 |
| Presence of Infection | 1.716 (1.231 – 2.394) | 0.002 |
| MASLD as etiology of cirrhosis | 0.906 (0.546 – 1.504) | 0.702 |

| *Table 6: Baseline characteristics of A-TANGO+ population, stratified by reversal status as per A-TANGO criteria* | | | |
| --- | --- | --- | --- |
| Characteristic | No reversal  (n=1150, 82.9%) | Reversal  (n= 238, 17.1%) | P-value |
| Age, median (IQR) | 46.0 (39.0 - 56.0) | 47.0 (38.0 - 53.0) | 0.564 |
| Female sex, n/N (%) | 108 (9.4) | 22 (9.2) | 1.000 |
| Presence of APASL ACLF | 512 (44.5) | 90 (37.8) | 0.067 |
| Etiology of cirrhosis (n, %) |  |  |  |
| Alcohol | 849 (73.8) | 184 (77.3) | 0.142 |
| MASLD | 130 (11.3) | 13 (5.5) |  |
| Hepatitis B/C | 35 (3.0) | 7 (2.9) |  |
| Alcohol+ CMRF | 20 (1.7) | 3 (1.3) |  |
| AIH | 71 (6.2) | 22 (9.2) |  |
| Cryptogenic | 10 (0.9) | 1 (0.4) |  |
| Alcohol+ Hepatitis B/C | 27 (2.3) | 7 (2.9) |  |
| Others | 8 (0.7) | 1 (0.4) |  |
| Precipitating events (n, %) |  |  |  |
| AH | 161 (14.0) | 41 (17.2) | 0.355 |
| Infections | 340 (29.6) | 53 (22.3) |  |
| AH+ others | 489 (42.5) | 104 (43.7) |  |
| DILI | 39 (3.4) | 10 (4.2) |  |
| Viral hepatitis | 23 (2.0) | 6 (2.5) |  |
| AIH | 37 (3.2) | 12 (5.0) |  |
| UGIB/LGIB | 38 (3.3) | 7 (2.9) |  |
| Others | 23 (2.0) | 5 (2.1) |  |
| Type of acute insult |  |  |  |
| Hepatic | 749 (65.1) | 173 (72.7) | 0.030 |
| Extra-hepatic | 401 (34.9) | 65 (27.3) |  |
| More than acute insult | 512 (44.5) | 109 (45.8) | 0.773 |
| Prior decompensation | 309 (37.6) | 40 (30.8) | 0.158 |
| Decompensations |  |  |  |
| Prevalence of Ascites | 1125 (97.8) | 231 (97.1) | 0.631 |
| Ascites grade |  |  |  |
| No Ascites | 25 (2.2) | 7 (2.9) | 0.663 |
| Mild to moderate | 636 (55.3) | 126 (52.9) |  |
| Tense ascites | 489 (42.5) | 105 (44.1) |  |
| Prevalence of HE | 996 (86.6) | 197 (82.8) | 0.148 |
| HE grades |  |  |  |
| No HE | 154 (13.4) | 41 (17.2) | <0.001 |
| Grade I | 294 (25.6) | 146 (61.3) |  |
| Grade II | 439 (38.2) | 18 (7.6) |  |
| Grade III | 107 (9.3) | 33 (13.9) |  |
| Grade IV | 156 (13.6) | 0 (0.0) |  |
| Infections | 384 (33.4) | 83 (34.9) | 0.715 |
| Lab investigations |  |  |  |
| Haemoglobin (gm/dL) | 9.1 (7.7 - 10.7) | 9.6 (8.1 - 11.5) | <0.001 |
| WBCC (x10^9^/L) | 11.6 (7.6 - 17.1) | 9.9 (7.0 - 15.3) | 0.002 |
| Platelet count (x10^9^/L) | 86.0 (54.0 - 137.0) | 101.5 (59.2 - 166.5) | 0.002 |
| INR | 2.3 (1.7 - 3.0) | 2.0 (1.3 - 2.5) | <0.001 |
| AST (U/L) | 104.3 (67.0 - 173.6) | 103.1 (60.2 - 161.3) | 0.466 |
| ALT (U/L) | 44.0 (29.0 - 82.9) | 41.7 (28.0 - 75.1) | 0.442 |
| Bilirubin (mg/dL) | 17.0 (7.1 - 26.0) | 16.0 (6.5 - 24.6) | 0.113 |
| Creatinine (mg/dL) | 1.6 (0.9 - 2.8) | 1.3 (0.9 - 2.4) | 0.016 |
| Sodium (mmol/L) | 131.0 (126.4 - 136.0) | 132.0 (127.9 - 136.0) | 0.180 |
| MAP (mmHg) | 84.0 (76.7 - 92.0) | 85.3 (80.0 - 93.3) | 0.009 |
| Disease severity scores |  |  |  |
| MELD score | 29.8 (25.0 - 35.7) | 27.0 (22.2 - 31.7) | <0.001 |
| MELD-Na score | 32.2 (27.9 - 36.9) | 29.7 (25.4 - 34.0) | <0.001 |
| A-TANGO OF score | 11.0 (10.0 - 13.0) | 10.0 (9.0 - 11.0) | <0.001 |
| AARC score | 11.0 (10.0 - 12.0) | 10.0 (9.0 - 11.0) | <0.001 |
| Organ failures |  |  |  |
| Cerebral failure | 263 (22.9) | 33 (13.9) | 0.003 |
| Coagulation failure | 649 (56.4) | 98 (41.2) | <0.001 |
| Liver failure | 499 (43.4) | 103 (43.3) | 1.000 |
| Kidney failure | 469 (40.8) | 85 (35.7) | 0.167 |
| Circulatory failure | 360 (31.3) | 38 (16.0) | <0.001 |
| Respiratory failure | 215 (18.7) | 12 (5.0) | <0.001 |
| 28-day mortality | 719 (62.5) | 22 (9.2) | <0.001 |
| 90-day mortality | 778 (67.7) | 42 (17.6) | <0.001 |
| INR: international normalized ration, AST: Aspartate Aminotransferase, ALT: Aspartate Aminotransferase, MAP: Mean arterial pressure, MELD: Model of end-stage liver disease, MELD-Na: MELD-sodium  Categorical variables are presented as n (%), and continuous variables as median (interquartile range, IQR). Categorical variables were compared using the Chi-square test, and continuous variables using the Mann–Whitney U test. A two-sided p-value <0.05 was considered statistically significant.  Complete follow up data till 28-days/death was available for 1388 patients out of 2715 A-TANGO+ patients at baseline | | | |

| *Table 7: Baseline characteristics of APASL+ population, stratified by reversal status as per APASL criteria* | | | |
| --- | --- | --- | --- |
| Characteristic | No reversal  (n=737, 96.0%) | Reversal  (n= 31, 4.0%) | P-value |
| Age, median (IQR) | 46.0 (40.0 - 54.0) | 49.5 (40.8 - 60.2) | 0.283 |
| Female sex, n/N (%) | 57 (7.7) | 3 (9.7) | 0.957 |
| Etiology of cirrhosis (n, %) |  |  |  |
| Alcohol | 582 (79.0) | 25 (80.6) | 0.236 |
| MASLD | 59 (8.0) | 0 (0.0) |  |
| Hepatitis B/C | 21 (2.8) | 3 (9.7) |  |
| Alcohol+ CMRF | 11 (1.5) | 0 (0.0) |  |
| AIH | 41 (5.6) | 3 (9.7) |  |
| Cryptogenic | 6 (0.8) | 0 (0.0) |  |
| Alcohol+ Hepatitis B/C | 13 (1.8) | 0 (0.0) |  |
| Others | 4 (0.5) | 0 (0.0) |  |
| Precipitating events (n, %) |  |  |  |
| AH | 131 (17.8) | 6 (19.4) | <0.001 |
| Infections | 192 (26.1) | 2 (6.5) |  |
| AH+ others | 310 (42.1) | 5 (16.1) |  |
| DILI | 34 (4.6) | 2 (6.5) |  |
| Viral hepatitis | 19 (2.6) | 2 (6.5) |  |
| AIH | 23 (3.1) | 2 (6.5) |  |
| UGIB/LGIB | 22 (3.0) | 12 (38.7) |  |
| Others | 6 (0.8) | 0 (0.0) |  |
| Type of acute insult |  |  |  |
| Hepatic | 517 (70.1) | 17 (54.8) | 0.106 |
| Extra-hepatic | 220 (29.9) | 14 (45.2) |  |
| More than acute insult | 321 (43.6) | 16 (51.6) | 0.483 |
| Prior decompensation | - | - | - |
| Decompensations |  |  |  |
| Prevalence of Ascites | 725 (98.4) | 31 (100.0) | 1.000 |
| Ascites grade |  |  |  |
| No Ascites | 12 (1.6) | 0 (0.0) | 0.828 |
| Mild to moderate | 415 (56.7) | 12 (60.0) |  |
| Tense ascites | 305 (41.7) | 8 (40.0) |  |
| Prevalence of HE |  |  |  |
| HE grades | 626 (84.9) | 18 (58.1) | <0.001 |
| No HE |  |  |  |
| Grade I | 108 (14.8) | 5 (25.0) | 0.007 |
| Grade II | 282 (38.5) | 14 (70.0) |  |
| Grade III | 242 (33.1) | 0 (0.0) |  |
| Grade IV | 59 (8.1) | 1 (5.0) |  |
| Infections | 41 (5.6) | 0 (0.0) |  |
| Lab investigations |  |  |  |
| Haemoglobin (gm/dL) | 9.7 (8.1 - 11.2) | 9.1 (7.2 - 11.6) | 0.149 |
| WBCC (x10^9^/L) | 11.2 (7.6 - 16.4) | 8.2 (5.7 - 9.0) | 0.002 |
| Platelet count (x10^9^/L) | 87.5 (54.0 - 149.0) | 91.0 (68.5 - 142.5) | 0.417 |
| INR | 2.3 (1.9 - 3.0) | 1.9 (1.7 - 2.7) | 0.024 |
| AST (U/L) | 112.0 (70.0 - 185.9) | 146.0 (110.5 - 274.3) | 0.027 |
| ALT (U/L) | 44.3 (29.0 - 88.0) | 75.5 (41.7 - 108.1) | 0.071 |
| Bilirubin (mg/dL) | 16.1 (9.0 - 24.6) | 8.3 (6.3 - 12.9) | <0.001 |
| Creatinine (mg/dL) | 1.5 (0.9 - 2.5) | 0.9 (0.8 - 1.8) | 0.047 |
| Sodium (mmol/L) | 130.4 (126.0 - 134.8) | 133.8 (129.8 - 138.2) | 0.014 |
| MAP (mmHg) | 83.3 (77.3 - 91.0) | 82.0 (75.0 - 87.5) | 0.249 |
| Disease severity scores |  |  |  |
| MELD score | 29.8 (25.4 - 36.6) | 25.0 (22.2 - 31.1) | 0.004 |
| MELD-Na score | 32.5 (28.4 - 37.4) | 30.1 (26.1 - 33.7) | 0.067 |
| A-TANGO OF score | 11.0 (9.0 - 13.0) | 9.0 (8.0 - 11.0) | 0.002 |
| AARC score | 11.0 (9.0 - 12.0) | 9.0 (8.0 - 9.8) | 0.001 |
| Organ failures |  |  |  |
| Cerebral failure | 100 (13.6) | 1 (3.2) | 0.162 |
| Coagulation failure | 407 (55.2) | 10 (32.3) | 0.020 |
| Liver failure | 288 (39.1) | 4 (12.9) | 0.006 |
| Kidney failure | 261 (35.4) | 7 (22.6) | 0.202 |
| Circulatory failure | 155 (21.0) | 5 (16.1) | 0.665 |
| Respiratory failure | 69 (9.4) | 3 (9.7) | 1.000 |
| 28-day mortality | 334 (45.3) | 3 (9.7) | <0.001 |
| 90-day mortality | 373 (50.6) | 5 (16.1) | <0.001 |
| INR: international normalized ration, AST: Aspartate Aminotransferase, ALT: Aspartate Aminotransferase, MAP: Mean arterial pressure, MELD: Model of end-stage liver disease, MELD-Na: MELD-sodium  Categorical variables are presented as n (%), and continuous variables as median (interquartile range, IQR). Categorical variables were compared using the Chi-square test, and continuous variables using the Mann–Whitney U test. A two-sided p-value <0.05 was considered statistically significant.  Complete follow up data till 28-days/death was available for 768 patients out of 2033 APASL+ patients at baseline | | | |

EASL ACLF analysis

| *Table 8: Baseline characteristics of cohort stratified by presence/absence of APASL* | | | | | | | |
| --- | --- | --- | --- | --- | --- | --- | --- |
|  | EASL- | | | EASL+ | | |  |
| Characteristic | APASL-  (n= 881, 21.9%) | APASL+  (n= 498, 12.4%) | P-value | APASL+  (n= 1532, 38.1%) | APASL-  (n= 1113, 27.7%) | P-value | Overall p-value |
| Age, median (IQR) | 48.0 (41.0 - 56.0) | 44.0 (37.0 - 52.0) | <0.001 | 45.0 (37.0 - 53.0) | 46.0 (38.0 - 55.0) | 0.002 | <0.001 |
| Female sex, n/N (%) | 92 (10.4) | 54 (10.8) | 0.888 | 177 (11.6) | 117 (10.5) | 0.436 | 0.794 |
| Etiology of cirrhosis (n, %) | | | | | | | |
| Alcohol | 639 (72.5) | 370 (74.3) | <0.001 | 1061 (69.3) | 803 (72.1) | <0.001 | <0.001 |
| MASLD | 67 (7.6) | 12 (2.4) |  | 112 (7.3) | 117 (10.5) |  |  |
| Hepatitis B/C | 75 (8.5) | 61 (12.2) |  | 183 (11.9) | 44 (4.0) |  |  |
| Alcohol+ CMRF | 23 (2.6) | 4 (0.8) |  | 16 (1.0) | 26 (2.3) |  |  |
| AIH | 29 (3.3) | 25 (5.0) |  | 66 (4.3) | 71 (6.4) |  |  |
| Cryptogenic | 25 (2.8) | 14 (2.8) |  | 70 (4.6) | 12 (1.1) |  |  |
| Alcohol+ Hepatitis B/C | 19 (2.2) | 11 (2.2) |  | 21 (1.4) | 35 (3.1) |  |  |
| Others | 4 (0.5) | 1 (0.2) |  | 3 (0.2) | 5 (0.4) |  |  |
| Precipitating events (n, %) | | | | | | | |
| AH | 93 (10.6) | 230 (46.2) | <0.001 | 613 (40.0) | 183 (16.4) | <0.001 | <0.001 |
| Infections | 137 (15.6) | 44 (8.8) |  | 190 (12.4) | 305 (27.4) |  |  |
| AH+ others | 177 (20.1) | 99 (19.9) |  | 303 (19.8) | 452 (40.6) |  |  |
| DILI | 33 (3.7) | 29 (5.8) |  | 123 (8.0) | 41 (3.7) |  |  |
| Viral hepatitis | 11 (1.2) | 50 (10.0) |  | 162 (10.6) | 27 (2.4) |  |  |
| AIH | 12 (1.4) | 4 (0.8) |  | 27 (1.8) | 32 (2.9) |  |  |
| UGIB/LGIB | 388 (44.0) | 15 (3.0) |  | 20 (1.3) | 52 (4.7) |  |  |
| Others | 30 (3.4) | 27 (5.4) |  | 94 (6.1) | 21 (1.9) |  |  |
| Type of acute insult | | | | | | | |
| Hepatic | 326 (37.0) | 412 (82.7) | <0.001 | 1228 (80.2) | 735 (66.0) | <0.001 | <0.001 |
| Extra-hepatic | 555 (63.0) | 86 (17.3) |  | 304 (19.8) | 378 (34.0) |  |  |
| More than acute insult | 410 (46.5) | 136 (27.3) | <0.001 | 403 (26.3) | 486 (43.7) | <0.001 | <0.001 |
| Decompensations | | | | | | | |
| Prevalence of Ascites | 651 (73.9) | 478 (96.0) | 0.698 | 1507 (98.4) | 1059 (95.1) | <0.001 | 0.001 |
| Ascites grade |  |  |  |  |  |  |  |
| No Ascites | 39 (7.6) | 20 (4.1) | <0.001 | 25 (1.6) | 52 (4.7) | <0.001 | <0.001 |
| Mild to moderate | 300 (58.3) | 362 (74.2) |  | 1132 (74.2) | 562 (51.1) |  |  |
| Tense ascites | 176 (34.2) | 106 (21.7) |  | 369 (24.2) | 485 (44.1) |  |  |
| Prevalence of HE | 352 (40.0) | 183 (36.7) | 0.264 | 1050 (68.5) | 939 (84.4) | <0.001 | <0.001 |
| HE grades |  |  |  |  |  |  |  |
| No HE | 169 (27.9) | 156 (45.9) | <0.001 | 630 (37.6) | 208 (20.7) | <0.001 | <0.001 |
| Grade I-II | 314 (51.9) | 148 (43.5) |  | 421 (25.1) | 251 (25.0) |  |  |
| Grade III-IV | 115 (19.0) | 36 (10.6) |  | 436 (26.0) | 298 (29.6) |  |  |
| Infection | 67 (61.5) | 170 (47.0) | 0.011 | 561 (57.0) | 468 (78.1) | <0.001 | <0.001 |
| Lab investigations | | | | | | | |
| Haemoglobin (gm/dL) | 8.6 (7.0 - 10.5) | 10.3 (8.7 - 11.6) | 0.001 | 9.9 (8.4 - 11.5) | 8.9 (7.4 - 10.7) | <0.001 | <0.001 |
| WBCC (x10^9^/L) | 7.4 (5.3 - 10.4) | 8.0 (5.9 - 12.2) | 0.004 | 12.0 (7.6 - 17.4) | 11.0 (7.1 - 16.5) | 0.002 | <0.001 |
| Platelet count (x10^9^/L) | 91.0 (69.0 - 130.0) | 99.0 (66.0 - 153.8) | 0.061 | 110.0 (69.0 - 174.0) | 88.0 (56.0 - 140.0) | <0.001 | <0.001 |
| INR | 1.4 (1.1 - 1.7) | 2.0 (1.7 - 2.2) | <0.001 | 2.3 (1.9 - 3.0) | 1.8 (1.2 - 2.5) | <0.001 | <0.001 |
| AST (U/L) | 73.1 (46.8 - 119.6) | 122.0 (78.8 - 197.2) | <0.001 | 140.0 (91.0 - 216.0) | 98.0 (63.0 - 156.6) | <0.001 | <0.001 |
| ALT (U/L) | 40.0 (23.7 - 61.8) | 52.0 (33.0 - 91.2) | <0.001 | 59.0 (35.0 - 114.0) | 42.0 (28.5 - 76.0) | <0.001 | <0.001 |
| Bilirubin (mg/dL) | 2.6 (1.3 - 5.0) | 9.0 (6.8 - 12.7) | <0.001 | 20.9 (14.2 - 27.4) | 14.0 (5.4 - 22.0) | <0.001 | <0.001 |
| Creatinine (mg/dL) | 0.9 (0.7 - 1.1) | 0.9 (0.7 - 1.1) | 0.022 | 1.5 (0.8 - 2.6) | 1.4 (0.9 - 2.5) | 0.832 | <0.001 |
| Sodium (mmol/L) | 134.8 (130.4 - 137.0) | 132.6 (128.2 - 136.0) | <0.001 | 131.0 (126.5 - 135.0) | 132.0 (128.0 - 137.0) | <0.001 | <0.001 |
| MAP (mmHg) | 80.0 (72.7 - 89.7) | 85.7 (78.0 - 92.0) | 0.230 | 83.3 (77.0 - 92.0) | 85.0 (77.3 - 93.3) | 0.061 | <0.001 |
| Disease severity scores | | | | | | | |
| MELD score | 14.5 (11.7 - 18.0) | 23.3 (21.4 - 25.5) | <0.001 | 31.2 (27.1 - 37.3) | 26.5 (19.7 - 32.0) | <0.001 | <0.001 |
| MELD-Na score | 19.3 (14.9 - 23.4) | 26.3 (23.5 - 28.9) | <0.001 | 33.2 (29.6 - 37.9) | 29.1 (23.8 - 34.1) | <0.001 | <0.001 |
| CLIF-C OF score | 7.0 (6.0 - 8.0) | 8.0 (8.0 - 9.0) | <0.001 | 11.0 (10.0 - 12.0) | 10.0 (9.0 - 12.0) | <0.001 | <0.001 |
| Organ failures | | | | | | | |
| Cerebral failure | 18 (2.0) | 3 (0.6) | 0.062 | 187 (12.2) | 238 (21.4) | <0.001 | <0.001 |
| Coagulation failure | 18 (2.0) | 18 (3.6) | 0.113 | 556 (36.3) | 420 (37.7) | 0.480 | <0.001 |
| Liver failure | 44 (5.0) | 134 (26.9) | <0.001 | 1261 (82.3) | 641 (57.6) | <0.001 | <0.001 |
| Kidney failure | 0 (0.0) | 0 (0.0) | - | 581 (37.9) | 402 (36.1) | 0.364 | <0.001 |
| Circulatory failure | 8 (0.9) | 13 (2.6) | 0.024 | 445 (29.0) | 332 (29.8) | 0.694 | <0.001 |
| Respiratory failure | 0 (0.0) | 21 (4.2) | <0.001 | 417 (27.2) | 135 (12.1) | <0.001 | <0.001 |
| 28-day mortality | 95 (10.8) | 87 (17.5) | 0.001 | 730 (47.7) | 596 (53.5) | 0.003 | <0.001 |
| 90-day mortality | 119 (13.5) | 128 (25.7) | <0.001 | 877 (57.2) | 656 (58.9) | 0.406 | <0.001 |
| Progression to EASL ACLF over 28-day follow up | 123/515 (23.9)* | 171/487 (35.1)* | <0.001 | NA | NA | - | - |
| Reversal of ACLF as per EASL | NA | NA | - | 44/548 (8.0)* | 161/895 (18.0)* | <0.001 | - |
| Reversal of ACLF as per APASL | NA | 14/215 (6.5)* | - | 17/553 (3.1)* | NA | - | 0.026 |
| INR: international normalized ration, AST: Aspartate Aminotransferase, ALT: Aspartate Aminotransferase, MAP: Mean arterial pressure, MELD: Model of end-stage liver disease, MELD-Na: MELD-sodium  Categorical variables are presented as n (%), and continuous variables as median (interquartile range, IQR). Categorical variables were compared using the Chi-square test, and continuous variables using the Mann–Whitney U test. A two-sided p-value <0.05 was considered statistically significant.  *Percentages calculated in patients in which complete follow-up data was available | | | | | | | |

| Table 9: Univariable Cox regression analysis for 28-day mortality across ACLF phenotypes as per EASL/APASL presence | | | |
| --- | --- | --- | --- |
| Phenotype | HR | 95% CI | p-value |
| APASL-/EASL- | Reference | - | - |
| APASL+/EASL- | 1.68 | 1.26 - 2.25 | <0.001 |
| APASL-/EASL+ | 6.66 | 5.36 - 8.27 | <0.001 |
| APASL+/EASL+ | 5.65 | 4.57 - 7.00 | <0.001 |

| Table 10: Predictors of progression to organ failures among EASL- population at baseline | | |
| --- | --- | --- |
|  | Univariable analysis | |
| Variable | sHR (95% CI) | p-value |
| Presence of ACLF-A | 1.578 (1.278 – 1.948) | <0.001 |
| CLIF-C OF | 1.480 (1.351 – 1.622) | <0.001 |
| Bilirubin | 1.016 (1.005 – 1.028) | 0.003 |
| INR | 1.158 (1.046 – 1.283) | 0.005 |
| Creatinine | 0.944 (0.828 – 1.076) | 0.386 |
| Sodium | 0.991 (0.989 – 0.993) | <0.001 |
| Albumin | 0.707 (0.594 – 0.842) | <0.001 |
| WBC | 1.012 (0.996 – 1.027) | 0.145 |
| Presence of Infection | 1.453 (1.163 – 1.816) | 0.001 |
| Presence of HE | 1.455 (1.182 – 1.792) | <0.001 |
| Multivariable analysis (Models adjusted for age, gender and etiology of cirrhosis) | | |
|  | Adj. sHR | p-value |
| Model 1 | | |
| Presence of ACLF-A | 1.268 (1.015–1.584) | 0.037 |
| CLIF-C OF score | 1.458 (1.317–1.615) | <0.001 |
| Model 2 | | |
| Presence of ACLF-A | 1.670 (1.355–2.057) | <0.001 |
| Presence of infection | 1.440 (1.151–1.803) | 0.001 |
| Model 3 | | |
| Presence of Infections | 1.334 (1.062–1.676) | 0.013 |
| CLIF-C OF score | 1.492 (1.357–1.641) | <0.001 |
| Model 4 | | |
| CLIF-C OF score | 1.507 (1.372–1.655) | <0.001 |
| WBC | 1.009 (0.993–1.025) | 0.264 |
| Model 5 | | |
| Presence of ACLF-A | 1.261 (1.009–1.576) | 0.042 |
| Presence of infection | 1.327 (1.056–1.667) | 0.015 |
| CLIF-C OF score | 1.436 (1.295–1.593) | <0.001 |

| Table 11: Predictors of reversal (EASL criteria) withing EASL+ population at baseline | | |
| --- | --- | --- |
|  | Univariable analysis | |
| Variable | sHR (95% CI) | p-value |
| Presence of APASL ACLF | 0.426 (0.306–0.593) | <0.001 |
| CLIF-C OF | 0.722 (0.661–0.788) | <0.001 |
| Bilirubin | 0.960 (0.947–0.973) | <0.001 |
| INR | 0.777 (0.671–0.900) | <0.001 |
| Creatinine | 0.886 (0.805–0.976) | 0.014 |
| WBC | 0.961 (0.939–0.984) | <0.001 |
| Platelet count | 1.002 (1.000–1.003) | 0.055 |
| Presence of HE | 0.350 (0.252–0.485) | <0.001 |
| Cerebral failure | 0.360 (0.218–0.593) | <0.001 |
| Liver failure | 0.497 (0.364–0.678) | <0.001 |
| CVS failure | 0.387 (0.257–0.583) | <0.001 |
| Multivariable analysis (Models adjusted for age, gender and etiology of cirrhosis) | | |
|  | Adj. sHR | p-value |
| Model 1 | | |
| Presence of APASL ACLF | 0.457 (0.329–0.635) | <0.001 |
| CLIF-C OF score | 0.732 (0.674–0.795) | <0.001 |
| Model 2 | | |
| Presence of APASL ACLF | 0.436 (0.314–0.605) | <0.001 |
| Presence of CVS failure | 0.419 (0.276–0.634) | <0.001 |
| Model 3 | | |
| CLIF-C OF score | 0.739 (0.679–0.805) | <0.001 |
| WBC | 0.977 (0.957–0.997) | 0.027 |
| Model 4 | | |
| Presence of APASL ACLF | 0.455 (0.328–0.631) | <0.001 |
| WBC | 0.970 (0.951–0.989) | 0.002 |
