## Supplementary figure 1 for "Harmonising APASL and A-TANGO criteria for acute-on-chronic liver failure: identification of complementary high-risk pre-ACLF populations": Figure S1.pdf

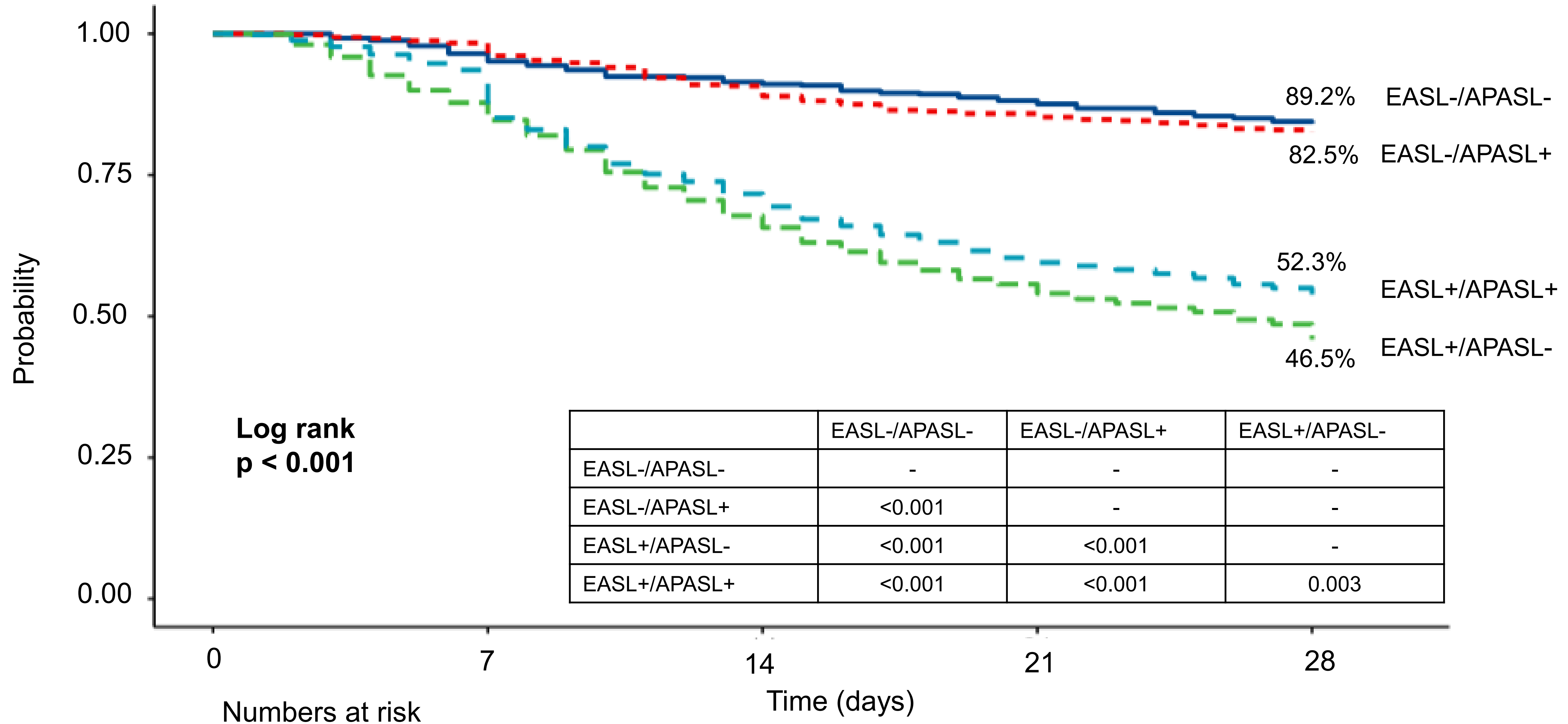


| | EASL-/APASL- | EASL-/APASL+ | EASL+/APASL- |
|---|---|---|---|
| EASL-/APASL- | - | - | - |
| EASL-/APASL+ | <0.001 | - | - |
| EASL+/APASL- | <0.001 | <0.001 | - |
| EASL+/APASL+ | <0.001 | <0.001 | 0.003 |

Numbers at risk

| | | | | | | |
|---|---|---|---|---|---|---|
| **EASL-** | **APASL-** | **515** | **589** | **553** | **527** | **508** |
| | **APASL+** | **340** | **328** | **306** | **292** | **289** |
| **EASL+** | **APASL-** | **1006** | **870** | **660** | **536** | **458** |
| | **APASL+** | **1677** | **1583** | **1234** | **1051** | **958** |
