## Supplementary figures and images for "Harmonising APASL and A-TANGO criteria for acute-on-chronic liver failure: identification of complementary high-risk pre-ACLF populations"

### Figure S2.pdf

#### Progression to organ failures over 28-days

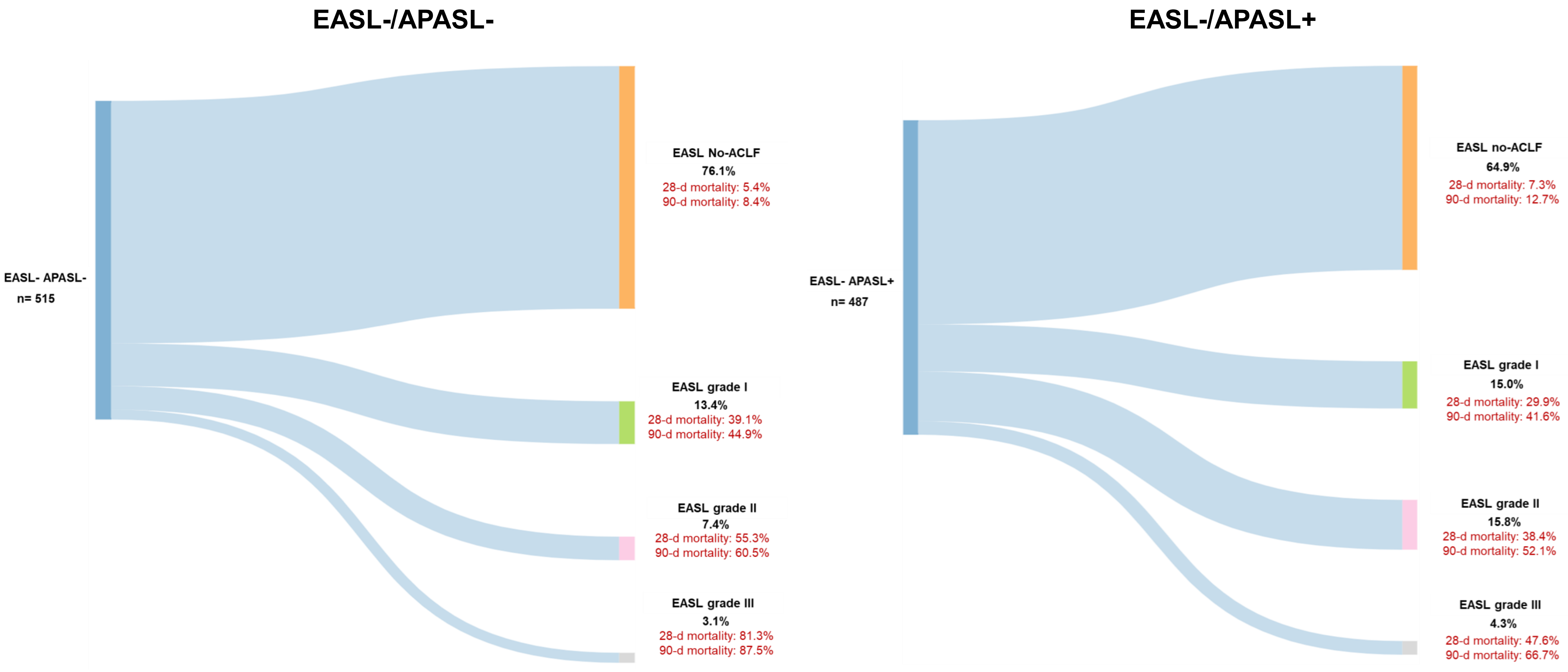
